# Long-read sequencing of an advanced cancer cohort resolves rearrangements, unravels haplotypes, and reveals methylation landscapes

**DOI:** 10.1101/2024.02.20.24302959

**Authors:** Kieran O’Neill, Erin Pleasance, Jeremy Fan, Vahid Akbari, Glenn Chang, Katherine Dixon, Veronika Csizmok, Signe MacLennan, Vanessa Porter, Andrew Galbraith, Cameron J. Grisdale, Luka Culibrk, John H. Dupuis, Richard Corbett, James Hopkins, Reanne Bowlby, Pawan Pandoh, Duane E. Smailus, Dean Cheng, Tina Wong, Connor Frey, Yaoqing Shen, Luis F. Paulin, Fritz J. Sedlazeck, Jessica M.T. Nelson, Eric Chuah, Karen L. Mungall, Richard A. Moore, Robin Coope, Andrew J. Mungall, Melissa K. McConechy, Laura M. Williamson, Kasmintan A. Schrader, Stephen Yip, Marco A. Marra, Janessa Laskin, Steven J.M. Jones

## Abstract

The Long-read POG dataset comprises a cohort of 189 patient tumours and 41 matched normal samples sequenced using the Oxford Nanopore Technologies PromethION platform. This dataset from the Personalized Oncogenomics (POG) program and the Marathon of Hope Cancer Centres Network includes accompanying DNA and RNA short-read sequence data, analytics, and clinical information. We show the potential of long-read sequencing for resolving complex cancer-related structural variants, viral integrations, and extrachromosomal circular DNA. Long-range phasing of variants facilitates the discovery of allelically differentially methylated regions (aDMRs) and allele-specific expression, including recurrent aDMRs in the cancer genes *RET* and *CDKN2A*. Germline promoter methylation in *MLH1* can be directly observed in Lynch syndrome. Promoter methylation in *BRCA1* and *RAD51C* is a likely driver behind patterns of homologous recombination deficiency where no driver mutation was found. This dataset demonstrates applications for long-read sequencing in precision medicine, and is available as a resource for developing analytical approaches using this technology.

## Main

Cancer is a multifaceted, heterogeneous disease that arises from a diverse array of genetic alterations. Comprehensive profiling methods have emerged as fundamental tools for deciphering the distinct genetic landscape and biology of each tumour and identifying therapeutic vulnerabilities^1,2^. While panel-based sequencing approaches have become routine in clinical settings^3,4^, the significance of whole genome and whole transcriptome analysis (WGTA) has progressively gained recognition, in both pediatric and adult cancers^5–8^. WGTA reveals driver mutations, gene fusions, expression alterations and genome signatures, significantly contributing to our understanding of cancer genome landscapes and informing tailored therapeutic choices for patients^9^.

Cancer profiling has to date been predominantly reliant on short-read sequencing methods, which while very successful have inherent challenges and constraints due to read length^10^. More recently, long-read sequencing, exemplified by Pacific Biosciences and Oxford Nanopore Technologies (ONT), can routinely produce reads of tens of thousands of bases, which impacts complex structural variant calling and ultra-long variant phasing^11^. In contrast, phasing to identify which variants occur on the same chromosome on the basis of short reads alone requires parental genotypes or statistical inference from reference populations. Another notable feature of sequencing native DNA using long-read technologies is the simultaneous detection of 5-methylcytosine^12,13^. Short-read methodologies require separate samples with an experimentally intensive assay for methylation detection (for example, bisulfite sequencing). DNA methylation is a key driver of many cancers, and characterizing DNA methylation has potential diagnostic, prognostic, and therapeutic applications^14,15^. Explorations of long-read sequencing in small cohorts of adult and pediatric tumours have proven fruitful, unveiling complex rearrangements, viral integrations, and tumour-specific methylation alterations^16–18^.

To achieve the potential of long-read sequencing in cancer genomics, the development of a comprehensive suite of analytical methods tailored explicitly for tumour analysis is imperative. Existing tools are often unsuited for cancer analysis or have been tested solely on cell-line data^19,20^. Patient-derived cancer samples encompass diverse features, including tumour heterogeneity, normal cell contamination from distinct tissues, and variability in mutation burden^1,2^, mandating analytical method refinement. To date, the absence of a sizable cohort of patient-derived cancer cases subjected to long-read sequencing has impeded progress in developing cancer-specific analytical approaches.

Here we present data from the Long-read POG cohort of 189 tumour samples obtained from 181 patients enrolled in the Personalized Oncogenomics (POG) program (NCT#02155621), sequenced using an ONT PromethION as part of the Marathon of Hope Cancer Centres Network. Each case in this cohort has also been studied using Illumina short-read normal whole genome sequencing (WGS), tumour WGS, and tumour RNA-seq. Our analyses illustrate the broad potential utility of this dataset in personalized oncogenomics. All data have been deposited in the European Genomics Archive as a resource for developers of software for tumour characterization from nanopore long-read data.

## Results

### The Long-read POG Cohort

Samples for long-read sequencing, previous short-read sequencing data, and short-read analysis were provided by the POG program, a precision medicine initiative that seeks to integrate WGTA into the clinical care of advanced cancer patients^2,6^ (Supplementary Table 1). The criteria used to select samples for long-read analysis included mutations in epigenetic modifiers, structural variant burden or homologous recombination deficiency (HRD), and presence of human papillomavirus (HPV). This Long-read POG cohort consists of 189 tumour samples from 181 patients. Of these, 43 tumour samples from 41 patients have matched normal nanopore sequencing allowing for somatic variant detection from long-read data. There were 26 cancer types represented, with the most common being breast (*n*=38, 20%), sarcoma (*n*=27, 14%) and colorectal (*n*=22, 12%) (Figure 1a). The majority of the tumours (*n*=144, 76%) were from biopsies of metastatic sites, frequently liver (*n*=63, 33%) and lymph nodes (*n*=30, 16%), while the rest were local recurrences or refractory disease (*n*=45, 24%). Patients received between zero and nine lines of systemic therapy before sequencing, and had a median overall survival of 34 months from diagnosis of advanced disease and 17 months from biopsy (Extended Data Figure 1a,b, Supplementary Table 1). The tumour mutation burden (TMB) determined from short-read variant analysis varied from 0.4 to 274 mutations per megabase (mut/Mb, median: 4.9), with seven cases exhibiting microsatellite instability (MSI; Figure 1b). HRD, as measured by the HRDetect score on short-read data, was considered high for 26 samples (14%), the majority of which (14/26) were breast or ovarian cancers. The tumour content ranged from 21-100% (median 67%).

**Figure 1.**
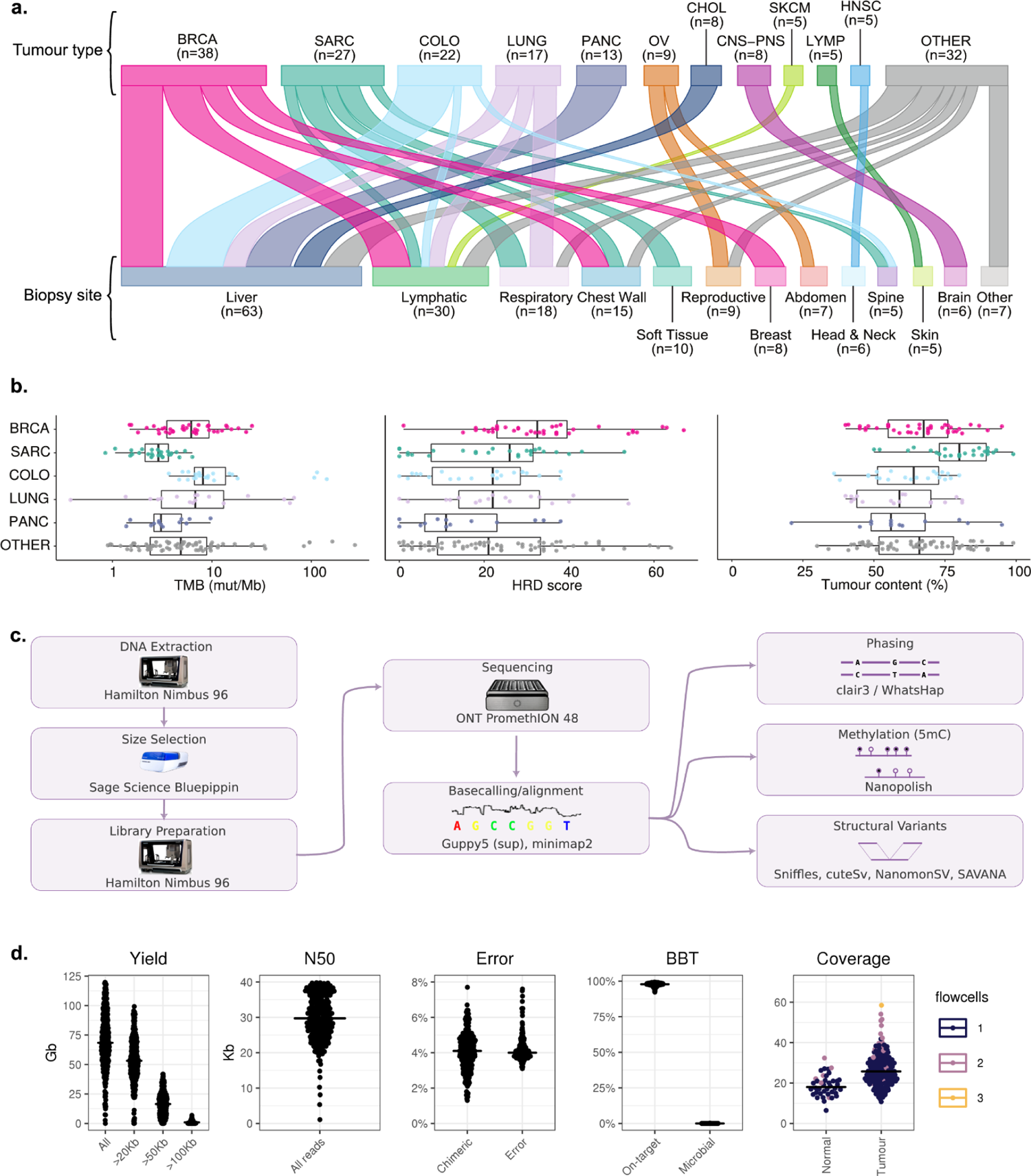
Nanopore long-read sequencing of an advanced cancer cohort. (a) Tumour types (top) and metastatic sites (bottom) for patient samples. Each patient is represented once; tissue groups with fewer than 5 samples are shown under ‘other’. (b) Genomic features of tumours by type. TMB, tumour mutation burden; mut/Mb, mutations per megabase; HRD, homologous recombination deficiency; BRCA, breast; SARC, sarcoma; COLO, colorectal; PANC, pancreatic. Boxplots represent the median, upper and lower quartiles of the distribution, and whiskers represent the limits of the distribution (1.5xIQR). ‘Other’ tumour group includes all tumours not in five most common tumour types, *n*=72. (c) Schematic overview of the laboratory methods and primary analysis for this cohort. (d) Approximate fold coverage per sample sequenced, and per-flowcell quality control statistics. Median yield of 68.4 Gbp per flowcell. Using BioBloom tools^21^, a median of 97.8% of reads matched the human reference, while no sample showed more than 0.2% of reads matching microbial taxa.

Automated library construction and nanopore sequencing on the PromethION platform (Figure 1c, Extended Data Figure 1c,d) yielded a median of 17.5-fold and 26-fold haploid genome coverage for normal and tumour samples, respectively (Figure 1d). The reads had a median N50 length of 31.3 kbp, with the longest read spanning 1,036,455 bp. Reads longer than 20 kb accounted for 77.8% of the sequence data. Median base error (the edit distance of aligned reads to the GRCh38 reference) was 4%. Chimeric artifacts were present in a median of 4.1% of the reads. Assessment for microbial contamination showed fewer than 0.2% of reads matching microbial taxa in any sample, which is below the false-discovery rate for the method used ^21^.

### Nanopore sequencing reveals novel complex SVs

We sought to evaluate the potential of nanopore sequencing, combined with currently available software, for detecting SVs in cancer samples. To this end, we applied four variant callers, two with the ability to call somatic events and two without (Figure 2a, 2b, Supplementary Table 3). We began by compiling a list of well-established oncogenic fusion events that were previously identified using short-read sequencing in this cancer cohort^22^. Of these, 8/8 were successfully identified in the nanopore sequencing data using a combination of SV callers (Supplementary Table 2), despite lower median sequencing depth of 26X for long-reads compared to short-reads typically at 80X.

**Figure 2.**
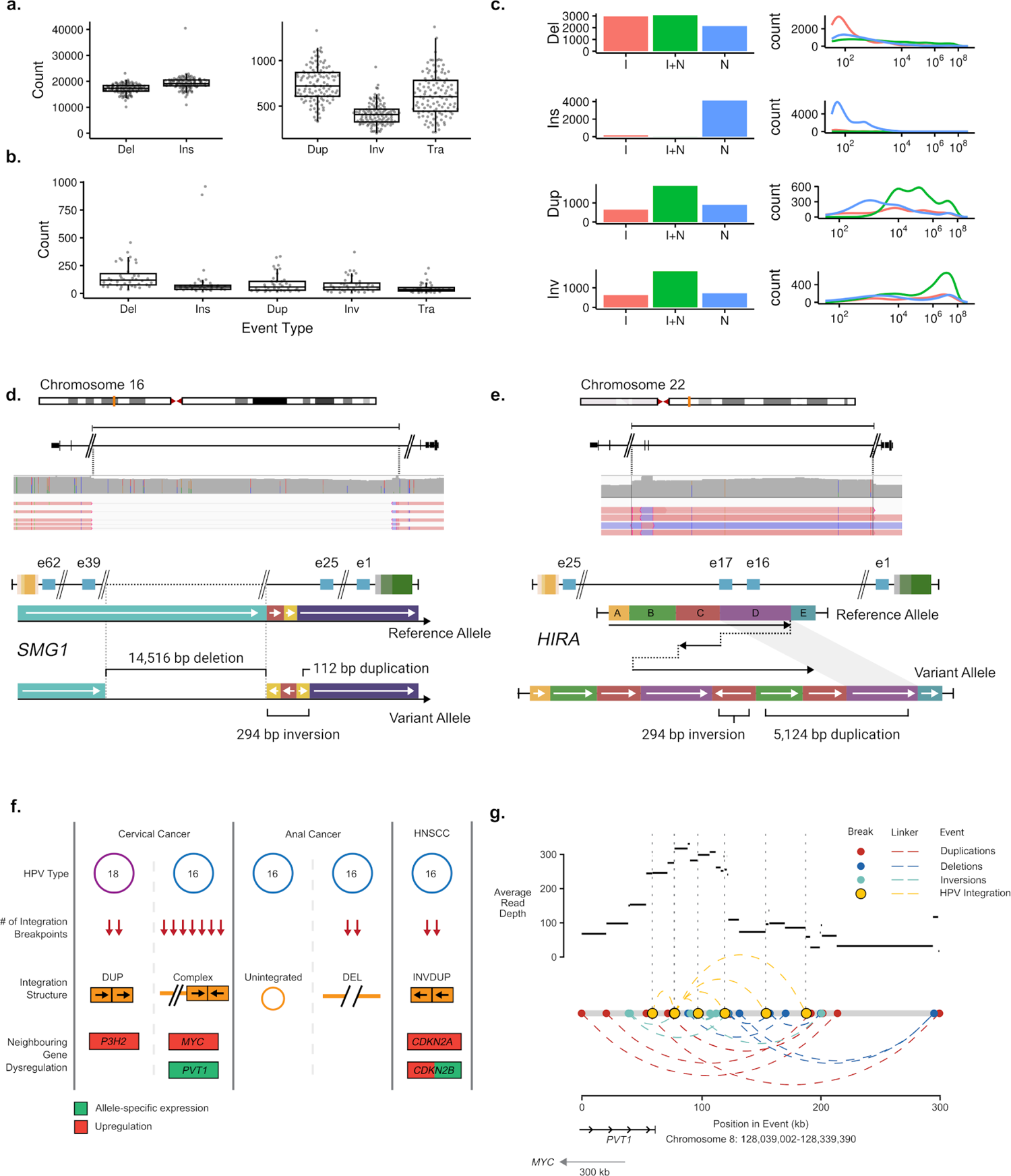
Structural Variants. (a) Per-sample counts of all SV calls (including germline) by type for all tumours (*n*=189). (b) Per-sample counts of somatic SV calls in samples with matched normal (*n*=43). (c) Concordance between high quality somatic Illumina SV calls (I) and nanopore SV calls with at least 1 caller support (N), summed across the cohort. (d) Schematic of a resolved complex foldback inversion affecting *SMG1*, including a deletion of exons 26-38, detected only in the nanopore data. (e) Schematic of a resolved complex foldback inversion affecting *HIRA*, including duplication of exon 16-17, detected only in the nanopore data. (f) Features of HPV integration characterised using nanopore sequencing in the five tumours with HPV. (g) Diagram of a complex rearrangement involving HPV integration sites in cervical cancer POG109.

We further compared somatic long-read calls with high-confidence somatic calls previously made in short-read data^2^ (see Methods). Of those, 1,919 (54.1%) duplications, 1,943 (57.1%) inversions, 3,358 (37.6%) deletions, and seven (<1.0%) insertions were consistent between the short- and long-read datasets (see Methods) (Figure 2c). To understand the disparities between the calls made on the different platforms, we manually reviewed those calls that overlapped cancer genes (Supplementary Table 5). The absence of calls in the nanopore results were attributed to lower coverage in 3/4 (75%) of samples. Conversely, all of the events unique to the nanopore calls (6/6) showed evidence in the underlying nanopore sequence alignment, but not in the short-read. In 4/6 samples this difference was attributed to low-complexity regions which could not be mapped with shorter reads. The remaining two calls were complex variants that could not be fully resolved by the short-read callers.

We examined these two complex SVs in depth. The first was a loss-of-function inversion, deletion and duplication in *SMG1* in a colorectal adenocarcinoma sample (POG117, Figure 2d). The second was an inversion with multiple duplication events that predicted a frameshift and likely loss of function of one allele of the tumour suppressor *HIRA* in a metastatic breast cancer sample (POG279, Figure 2e). These complex SVs showed an overlapping breakpoint in an L1MB4 LINE element and an AluY element respectively, underscoring the capacity of long-read sequencing to resolve some SVs in repetitive regions.

The greatest disparity in somatic calls was a notably greater number of insertions called in the nanopore data (Figure 2c). We manually reviewed the underlying sequence data for those affecting cancer genes. Of these, 8/14 were identified as miscalls resulting from difficult-to-map regions. The remaining 6/14 had underlying nanopore sequence evidence but were missed or mischaracterized by short-read callers (Supplementary Table 4; Supplementary Figure 1a,b). Notably, all 14 of these insertion calls were made by a single SV caller, nanomonSV. This highlights the potential for long reads but also the need for further development of nanopore somatic SV calling software.

Four samples exhibited clear outlier behaviour in terms of the number of SV calls of a particular type (Extended Data Figure 2a). Two patients, POG884 and POG986, had tenfold more insertion calls than the median, but only in the nanopore calls, accounting for 1,837/4,126 of the Nanopore-only insertion calls mentioned above. These were the only two cases (of those with somatic calls) with MSI. On examining the insertions themselves, the majority (71.5%) overlapped short tandem repeat regions, consistent with MSI repeat expansions (Extended Data Figure 2b)^23^. Two other cases, POG111 and POG147, exhibited high nanopore-unique inversion frequencies. These showed copy number alteration (CNA) and inversion profiles which have been previously characterised as “tyfonas”^24^ (Extended Data Figure 3a,b,c).

### Oncoviral integrations detected by long reads impact surrounding gene expression

Human papillomavirus (HPV) infection is the driving cause of cervical cancer and implicated in many head and neck and anogenital cancers. HPV integration into the host genome is frequent, and integration events often involve a complex combination of structural alterations and multiple copies of the 8 kb viral genome. This complexity makes mapping with short reads difficult. We investigated the ability of long-read sequencing to reconstruct HPV integration events and their effects.

We investigated five samples with HPV previously detected in short-read data and confirmed in this study. Of these, four had HPV integration detected, each at a single host genome location (Figure 2f, Supplementary Table 1). We identified three of these events as simple, meaning that they were made up of only two integration breakpoints. The remaining event, in POG109, was complex, incorporating seven HPV-to-host genome breakpoints within a 130 kb region in the 8q24 locus, 300 kb downstream (3’) of *MYC* and overlapping the *MYC*-associated lncRNA *PVT1* (Figure 2g). This event also overlapped several SVs and oscillating copy number states, which resembled focal chromothripsis (Extended Data Figure 4b).

Three of the integration events were associated with elevated expression (≥85th percentile^2^; Extended Data Figure 4a) of neighbouring cancer genes, including *P3H2*, *MYC*, *CDKN2A* and *CDKN2B*. POG109 exhibited allele-specific expression (ASE) of *PVT1*, the gene it overlapped, with higher expression from the haplotype containing the integration. POG785 (which had overexpression of *CDKN2A* and *CDKN2B*), exhibited ASE of *CDKN2B*, also with higher expression from the haplotype containing the integration.

### Long-range phasing enables resolution of double hits to tumour suppressors

We assessed the ability of nanopore sequencing to enable long-range phasing, particularly of tumour suppressor genes (TSGs). The biallelic inactivation of TSGs is an important mechanism of tumour formation, with potential biological and clinical significance for informing diagnosis, disease prognosis and/or response to therapy. We found that phase block size was strongly correlated with read length (Spearman’s rho 0.72, *P* ≤ 2.2×10^-16^, Figure 3a). Phasing was able to completely resolve the haplotypes of the majority of genes in each sample, from promoter to 3’ end (median 85%, IQR 79.1-89.1%). This included most putative and known tumour suppressor genes (Figure 3b), although longer genes were completely phased less often (Spearman’s rho −0.65, *P* ≤ 2.2×10^-16^). However, several notable tumour suppressors of modest size, including *BRCA1*, *NF1* and *RB1*, could only be phased completely in around half of tumours, suggesting other locus-related features may reduce their phasing potential. Further investigation showed that this was due to lower density of phasing SNPs and greater density of repetitive or paralogous sequences that may not be fully resolved at modest read lengths (Extended Data Figure 5). In tumour genomes, ploidy, genomic instability, loss of heterozygosity and somatic variation may further influence global and local phasing.

**Figure 3.**
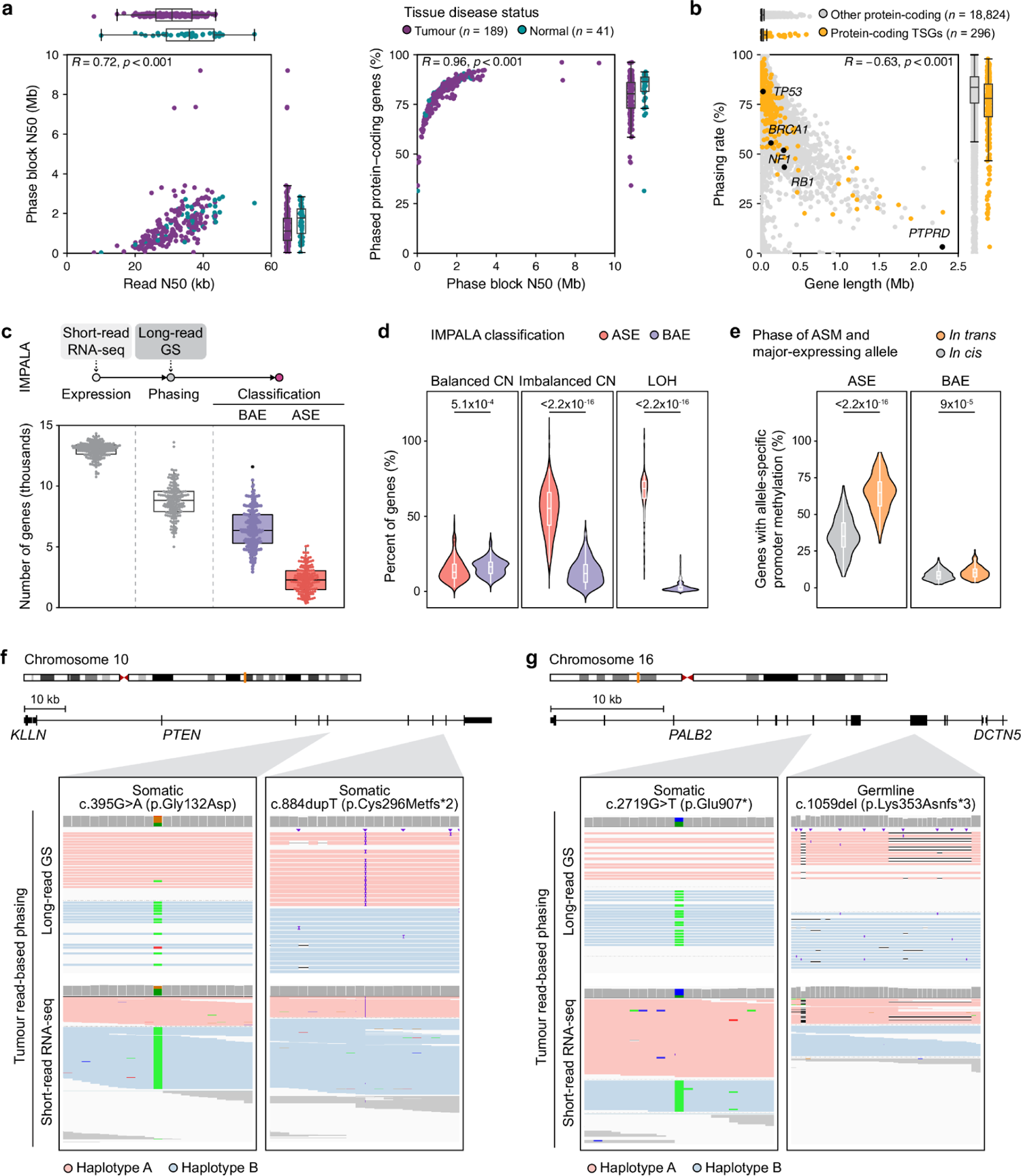
Phasing. (a) Correlation between read length, phase block size and phasing rate for Ensembl 100 protein-coding genes (plus promoters) across normal and tumour tissues. (b) Correlation between gene length and phasing rate for protein-coding genes (percent of tumours in which a gene plus promoter could be fully phased). (c) Summary of IMPALA results for the cohort, showing number of genes with sufficient expression to be considered (<1 TPM), number with sufficient expression and at least one phasing SNP, and their final classification as having allele-specific expression (ASE) or balanced allelic expression (BAE). (d). Percent of genes in regions of the tumour genome with balanced copy number (CN), imbalanced CN, or LOH that were classified as ASE or BAE. (e) Percent of genes with allele-specific promoter methylation by the relative phase of the major expressed allele for ASE and BAE genes. Examples of biallelic variants in tumour suppressor genes with ASE (f) and BAE (g). Reads are coloured by predicted haplotype from long read-based phasing, and reads that could not be assigned to a haplotype are coloured in grey.

Biallelic tumour suppressor inactivation may occur via different combinations of loss of heterozyosity (LOH) and small mutations. When two small inactivating mutations are called by short-read sequencing in the same TSG, it is often assumed that they are *in trans*. In the Long-read POG cohort, cases with two or more small somatic variants in tumour suppressor genes with potential biological and/or clinical significance were identified by retrospective review of the genomic report issued at the time of POG analysis. There were 30 cases identified with double somatic variants in at least one tumour suppressor gene. Among 33 pairs of variants (with three variants in one case), 19 pairs across 18 cases could be phased by long reads (Table 1). Variants for which phase could not be confirmed included variants supported by only one read, variants not within a phase block, and variants supported by reads with conflicting haplotype assignments. The majority (n = 17) of pairs were found to occur *in trans*, while two were found to occur *in cis*: double somatic variants in *PTEN* (POG507) and *KAT5* (POG446). Notably, the double somatic variants in *PTEN* were found to occur on the opposite allele from a heterozygous somatic *PTEN* deletion, suggesting an alternate mechanism of biallelic loss.

**Table 1:** Phasing of tumour suppressor gene small variants.

| Phase | Origin | Case ID | Cancer | Gene | Allele A | Allele B |
| --- | --- | --- | --- | --- | --- | --- |
| *In trans* | Germline & somatic | POG1000 | PANC | *ATM* | Exon 9 deletion (germline) | c.3577-14_3585delinsC (somatic) |
|  |  | POG792 | CHOL | *BRCA2* | p.(Leu2092Profs*7) (germline) | p.(Val2385Phefs*9) (somatic) |
|  |  | POG976 | CHOL | *PALB2* | p.(Lys353Asnfs*3) (germline) | p.(Glu907*) (somatic) |
|  |  | POG604 | BRCA | *TP53* | p.(Arg213*) (germline) | p.(Glu180*) (somatic) |
|  | Double somatic | POG295 | COLO | *APC* | p.(Cys417Valfs*37) | p.(Tyr1376Cysfs*9) |
|  |  | POG720 | COLO | *APC* | p.(Ala591Profs*19) | p.(Pro1427Lysfs*44) |
|  |  | POG777 | OV | *ARID1A* | p.(Gln1401*) | p.(Met1595Val) |
|  |  | POG446 | BRCA | *KAT5* | p.(Ile37Met) & p.(Ser135Phe) |  |
|  |  | POG130 | COLO | *MLH1* | p.(Glu297*) | p.(Phe530Serfs*5) |
|  |  | POG581 | SKCM | *NF1* | p.(Ser879*) | p.(Gln1188His) |
|  |  | POG239 | SECR | *NOTCH1* | p.(Ala1349Leufs*53) | p.(Cys942Tyr) |
|  |  | POG777 | OV | *PIK3R1* | p.(Thr371Ilefs*5) | p.(Lys459del) |
|  |  | POG352 | UCEC | *PTEN* | p.(Arg130Gly) | c.1026+1G>T |
|  |  | POG778 | BRCA | *PTEN* | p.(Tyr27Asp) | p.(Leu247Phefs*6) |
|  |  | POG958 | BRCA | *PTEN* | p.(Ile32Asn) & p.(Ala34Gly) | c.490_492+1del |
|  |  | POG884 | ESCA | *RB1* | p.(Arg255*) | p.(Met484Valfs*8) |
|  |  | POG021 | LUNG | *TP53* | p.(Val272Leu) | p.(Gly154Val) |
|  |  | POG680 | HNSC | *TP53* | p.(Pro82Leu) & p.(Ser127Pro) | p.(Arg282Trp) |
| *In cis* | Double somatic | POG446 | BRCA | *KAT5* | p.(Ile37Met) & p.(Ser135Phe) |  |
|  |  | POG507* | BRCA | *PTEN* | p.(Pro38Ser) & p.(Phe278Leu) |  |

\*Double somatic variants occurred in the context of copy loss of the other allele, consistent with biallelic events in *PTEN*.

### Long-range variant phasing facilitates the detection of allele-specific expression (ASE) and linkage to genomic events

ASE is an imbalance in expression between alleles of a gene, which is typically mediated by CNA or cis-acting regulatory mechanisms^25,26^. Long-range phasing offers the potential to more accurately determine ASE and link it to genomic lesions within the same or nearby genes^16,27^. We used the IMPALA pipeline to examine ASE in the Long-read POG cohort.

We found ASE in an average of 26.5% of the phased genes within each sample (SD = 12.2%) (Figure 3c). CNAs have been identified as the primary drivers of ASE genes in cancer cells^28,29^, and our results recapitulated this. Within this cohort, ASE genes tended to overlap regions of LOH (*P*=6.3×10^-115^) and copy number imbalance (*P*=1.1×10^-65^) whereas genes with biallelic expression (BAE) overlapped copy number balanced regions (*P*=0.02) (Figure 3d). We further noted a significant positive correlation between the CNA allelic ratio and the major expressed allele frequency (*r*=0.63, *P*≤2.2×10^-16^). ASE may also be due to epigenomic dysregulation. Examining ASE in regions with balanced copy number, we found a significantly greater proportion of genes with promoter allelic methylation (mean=0.16) than BAE (mean=0.01) (*P*=3.5×10^-23^). Moreover, allelic promoter methylation was more commonly found *in trans* with the major expressed allele (*P*≤2.2×10^-16^) (Figure 3e), indicating lower expression of the methylated allele.

ASE can be used to validate the downstream expression effects of aberrant cis-acting regulatory mechanisms and further ascertain biallelic TSG inactivation. For example, *PTEN* in POG041 shows ASE with major expressed allele frequency of 0.67. A frameshift and missense mutation (rs121909241) were found on the minor and major expressed allele respectively, which represents a double hit knockout scenario consistent with the gene’s ASE (Figure 3f). ASE was also seen in POG976 with *PALB2* somatic and germline variants, confirmed by phasing to be opposite alleles. Consistent with loss of function of *PALB2*, this cholangiocarcinoma was characterised by strong mutational signatures of HRD (Figure 3g).

The most frequent ASE gene in this cohort was *DUSP22*, in 122/135 samples in which it was expressed and could be phased, with a median major expressed allele frequency of 0.95. *DUSP22* expression is associated with poorer survival in low-grade lymphomas^30,31^. It also shows tissue-specific imprinting during brain development^32,33^. A survey of normal tissues from GTEx showed that only 9.12% (542/5940) of *DUSP22*-expressing samples showed ASE^34^. In the Long-read POG cohort, 63.11% of tumour samples with ASE in *DUSP22* showed allele-specific promoter methylation *in trans* and 81.15% showed allele-specific gene body methylation *in cis* with the major expressed allele (Extended Data Figure 6a). Although allele-specific promoter methylation is found in both blood and tumour samples, allelic loss of gene body methylation is a tumour specific phenomenon (*P*≤2.2×10^-16^, Extended Data Figure 6a-c). This suggests that reactivation of tissue-specific imprinting of *DUSP22* may play a role in tumourigenesis.

### DNA methylation derived from nanopore sequencing can reveal global methylation patterns and reflect tissue of origin

We evaluated nanopore-derived DNA methylation calls within this study for their potential in personalized oncogenomics. A comparison of DNA methylation detected using nanopore sequencing with whole genome bisulfite sequencing (WGBS) calls from the same sample showed good correlation (*R*=0.87, Figure 4a). In the Long-read POG cohort, tumours displayed significant hypomethylation compared to normal WGBS data from best-match tissue types (Figure 4b; see Methods). This hypomethylation was most distinct in repetitive regions. Notably, these regions are more readily mappable with long-read alignment^35^. The only genomic regions with hypermethylation in tumour samples compared to normal WGBS data (P=1.4×10^-5^) were CpG islands (CGIs). These results are consistent with the previously-described pattern of overall hypomethylation but focal hypermethylation in tumour DNA^36^.

**Figure 4.**
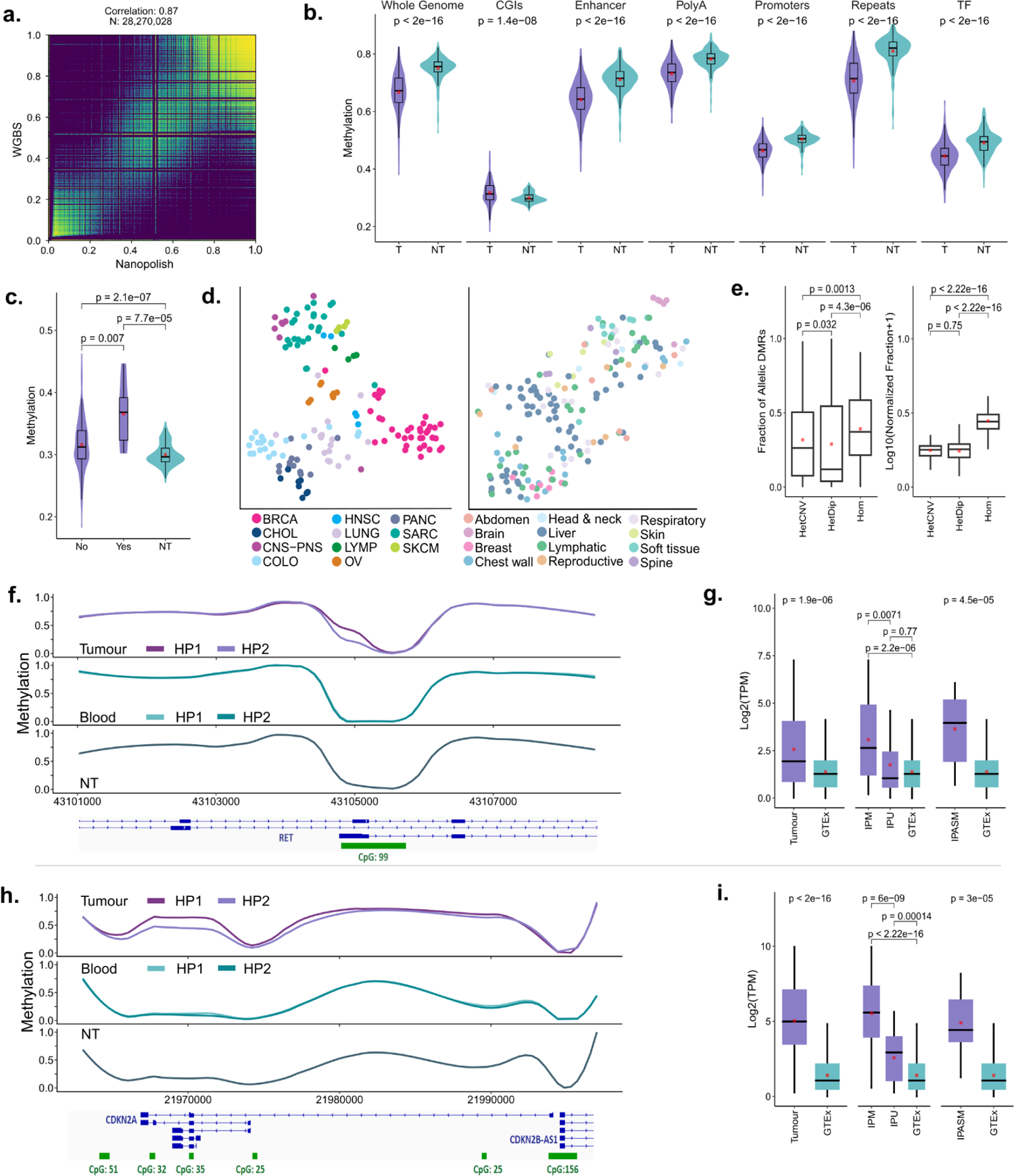
Methylation. (a) Correlation of nanopolish methylation frequency with WGBS for POG044. (b) Average methylation across tumours (T) compared with public WGBS methylation data from normal tissues and cells (NT), genome-wide and at different genomic regions. (c) Average methylation at CGIs in POG cases with iether IDH activating or TET-inactivating mutations (Yes) compared with the remainder of the cohort (No) and public normal tissue (NT). (d) tSNE plots based on DNA methylation at regulatory regions, compared with tumour type (left) and biopsy site (right). (e) aDMR distributions by copy number (CN). HetDip indicates CN balanced regions. HetCNV indicates CNV regions with both parental alleles. Hom indicates LOH. (f) DNA methylation at *RET* intragenic promoter CpGs, compared with patient blood and tissue normals (NT). (g) *RET* gene expression compared with GTEx normal tissues in: Left: the whole cohort. Centre: samples with >25% intragenic promoter methylation (IPM) vs other samples (IUM). Right: only those samples with an aDMR at the intragenic promoter (IPASM). Only samples with TPM>1 were used for expression comparison. (h-i) The same analysis as (f-g) but for *CDKN2A*. Note that in f and h the haplotags were swapped so that HP1 represents the hypermethylated allele. All P-Values are Wilcoxon rank-sum test.

TET enzymes are involved in active DNA demethylation, and use α-ketoglutarate as a cofactor, which is a product of IDH enzyme activity. TET and IDH mutations are recurrent in cancer^37^ and can result in hypermethylation of tumour genomes. Within the cohort, 10/189 samples (18/181 cases) had IDH gain of function mutations or TET candidate inactivating mutations detected using short-read sequencing (Supplementary Table 1). Compared with other cases and normal tissue samples, cases with mutations in IDH and TET genes show similar methylation patterns at all regulatory sites except for CGIs (Figure 4c). At CGIs, mutated samples show slight hypermethylation compared to the rest of tumour samples and WGBS normal samples. TET enzymes demonstrate sequence specificity toward CGIs^37–39^. The slightly higher methylation only at CGIs in the mutated samples in our cohort highlights the sequence specificity of TET enzymes and suggests that the genome-wide hypomethylation in tumour samples is largely due to the passive DNA demethylation pathway^40–42^.

Methylation patterns can distinguish tissue and tumour type^43,44^. We performed tSNE analysis of methylation in this cohort as a coarse assessment of this. We observed that samples tended to group by tumour tissue of origin, irrespective of metastatic biopsy site (Figure 4d). This finding suggests the potential utility of nanopore-derived DNA methylation for detecting or confirming tissue of origin in advanced and metastatic cancers, as an adjunct or complimentary analysis to the RNA approaches currently in use^45^.

### Patterns of allele-specific methylation in promoters and gene bodies are uncovered by long-range phasing

As shown earlier in this study and elsewhere, long reads enable long-range phasing, which extends to methylation information^46^. In this cohort, an average of 84% of the CpG sites within each sample could be phased (median= 86%; SD= 7.1%). We define the term “allelically differentially methylated regions” (aDMRs) to refer to genomic regions in which clusters of CpG sites display differential methylation between alleles. We detected 4.46 million (mean=23.61K, SD=14.64K) aDMRs across all tumour samples, with around five-fold more in tumour samples than their matched normals (mean=4.7K, SD=876; Extended Data Figure 7a). The majority (79%) of the aDMRs mapped to het-CNV and LOH regions (Figure 4e). The number of aDMRs in each sample was positively correlated with the fraction of the genome in LOH regions (*r*=0.469, *P*=2.6×10^-11^), and negatively correlated with the fraction of the genome in het-diploid regions (*r*= −0.38, *P*=1.4×10^-7^).

We examined aDMRs in het-diploid regions as potential sources of cancer-specific epigenetic dysregulation. We excluded aDMRs associated with normal cell function (linked to imprinting or random allele-specific methylation; see Methods), leaving ∼462K tumour-specific aDMRs. Most (76% of aDMRs) mapped to CGI, TF, promoter, enhancer or polyA sites (Extended Data Figure 7b). We detected 2,854 genes (8,511 transcripts) with recurrent aDMRs at their promoters, including the cancer genes *RET* and *CDKN2A*. Transcription factor (TF) enrichment analysis of these genes demonstrated significant enrichment for PRC1 and PRC2 complex protein subunits (adjusted *P*<0.05, Extended Data Figure 7c).

We further examined the aDMRs affecting *RET* and *CDKN2A*. *RET* is a receptor tyrosine kinase and a well-known oncogene^47^. The *RET* aDMR overlapped an intragenic CGI alternate promoter (per Ensembl 100 transcript models) in 26 of the cohort samples (Figure 4f). Another 66 tumour samples showed methylation (>25% average methylation) at this promoter. Those cases with intragenic promoter methylation (allelic or non-allelic) had significantly greater overall *RET* gene expression than those without methylation (*P*=0.0071), and normal GTEx tissues (*P*=2.2×10^-6^, Figure 4g).

*CDKN2A* is a well-known tumour suppressor gene that has a series of intragenic CGI promoters, which showed recurrent aDMRs in nine samples (Figure 4g) and methylation (>25% average methylation) in 140 samples. As for *RET*, cases with methylation (allelic or non-allelic) at this intragenic promoter had significantly greater expression of *CDKN2A* compared to those without methylation (*P*=6×10^-9^) and normal GTEx data (*P*<2.22×10^-16^, Figure 4i). Despite *CDKN2A* being a tumour suppressor, it is sometimes upregulated in cancers, with associated increase in immune cell infiltration^48^, (Extended Data Figure 8b), suggesting that it may function as an oncogene. In both *RET* and *CDKN2A,* intragenic promoter methylation may be a novel means of inducing overexpression of these genes as part of tumourigenesis.

### Epigenetic inactivation of DNA repair genes

HRD is especially prevalent in breast and ovarian cancers^49^, and its presence is relevant for therapeutic selection. HRD can arise due to inactivation of DNA repair genes by a combination of mutations and promoter methylation. We evaluated the promoter methylation frequencies of 51 homologous recombination (HR) genes in a combined breast (n=40) and ovarian (n=8) cohort (see Methods). Three breast cancer samples showed *BRCA1* promoter hypermethylation (Table 2). *RAD51C* promoter hypermethylation was observed in one breast and two ovarian cancer samples (Table 2). All six samples exhibited high HRDetect scores (≥0.7; see Methods) consistent with an HRD phenotype (Figure 5a). No deleterious somatic or pathogenic germline mutations were found in five HR genes (*BRCA1*, *BRCA2*, *ATM*, *PALB2*, and *RAD51C*) in these samples, suggesting that silencing of *BRCA1* and *RAD51C* was likely the primary cause of the observed HRD (Figure 5a).

**Figure 5.**
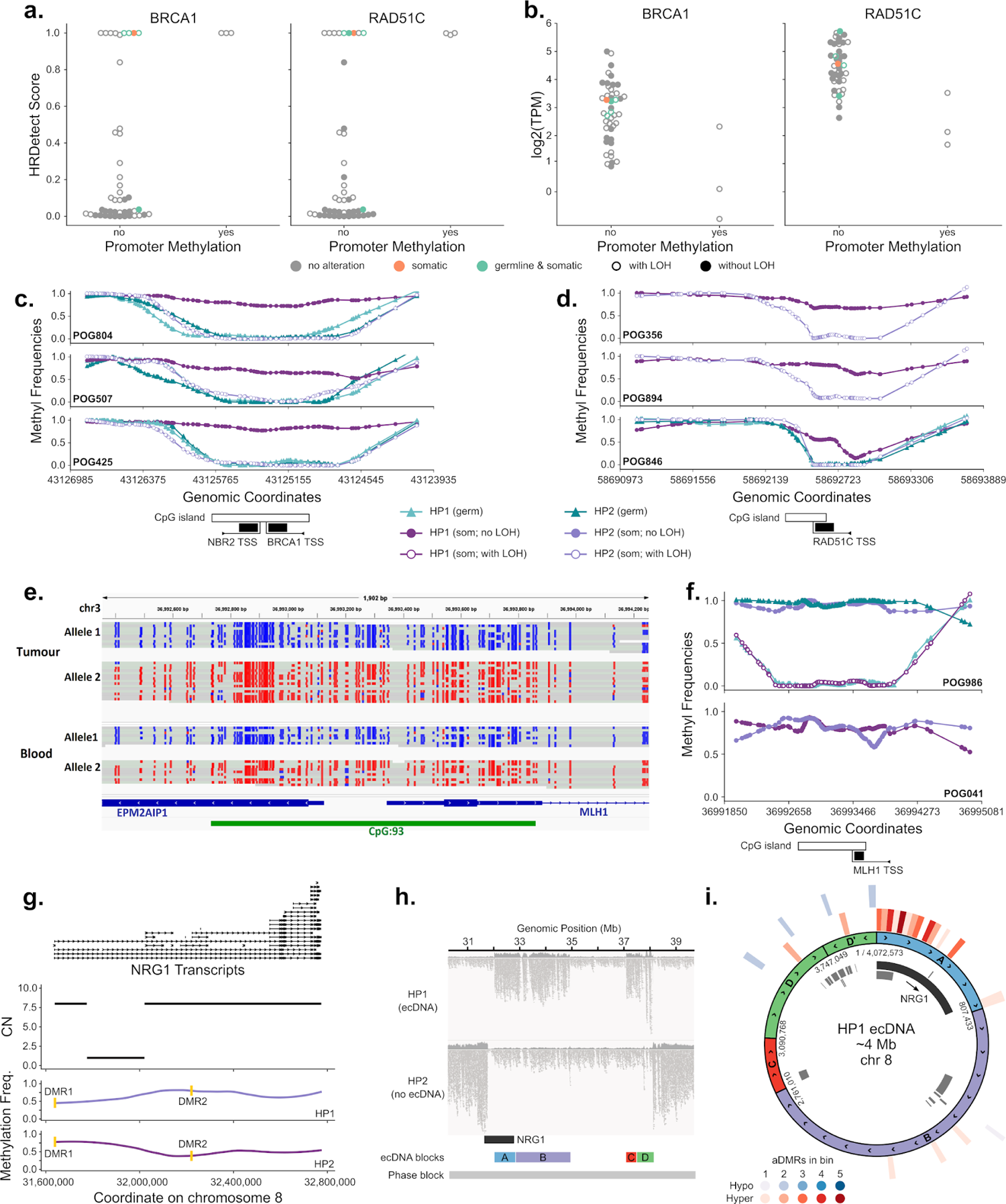
Integrative analyses. HRDetect scores (a) and expression values (b) for breast and ovarian samples with or without promoter methylation in *BRCA1* or *RAD51C*. Samples with deleterious alterations in five key HR genes (*BRCA1, BRCA2, ATM, PALB2, RAD51C*) are coloured. Haplotype-specific DNA methylation frequencies at the *BRCA1*/*NBR2* (c) and *RAD51C* (d) promoter regions in HRD samples (HRDetect score>=0.7) with promoter methylation. (e) Haplotype-specific DNA methylation at the *MLH1* promoter in a lung squamous cell carcinoma sample with *MLH1* germline epimutation. (f) Haplotype-specific DNA methylation frequencies at the *MLH1* promoter region in a lung squamous cell carcinoma sample with *MLH1* germline epimutation (top) and in a uterine endometrioid carcinoma sample with somatic *MLH1* promoter methylation (bottom). (g) Haplotype-specific methylation and copy number for *NRG1* in breast cancer sample POG816. The 3’ amplification was included within an ecDNA. Promoter aDMRs are highlighted in yellow. (h) Haplotype-phased long reads mapped to the ecDNA region. (i) Circos plot of the *NRG1*-containing ecDNA, highlighting DMRs and methylation states. Inner track: gene annotations, with NRG1 highlighted. Outer tracks: binned counts of aDMRs, showing substantial enrichment at the 5’ end of NRG1.

**Table 2:**
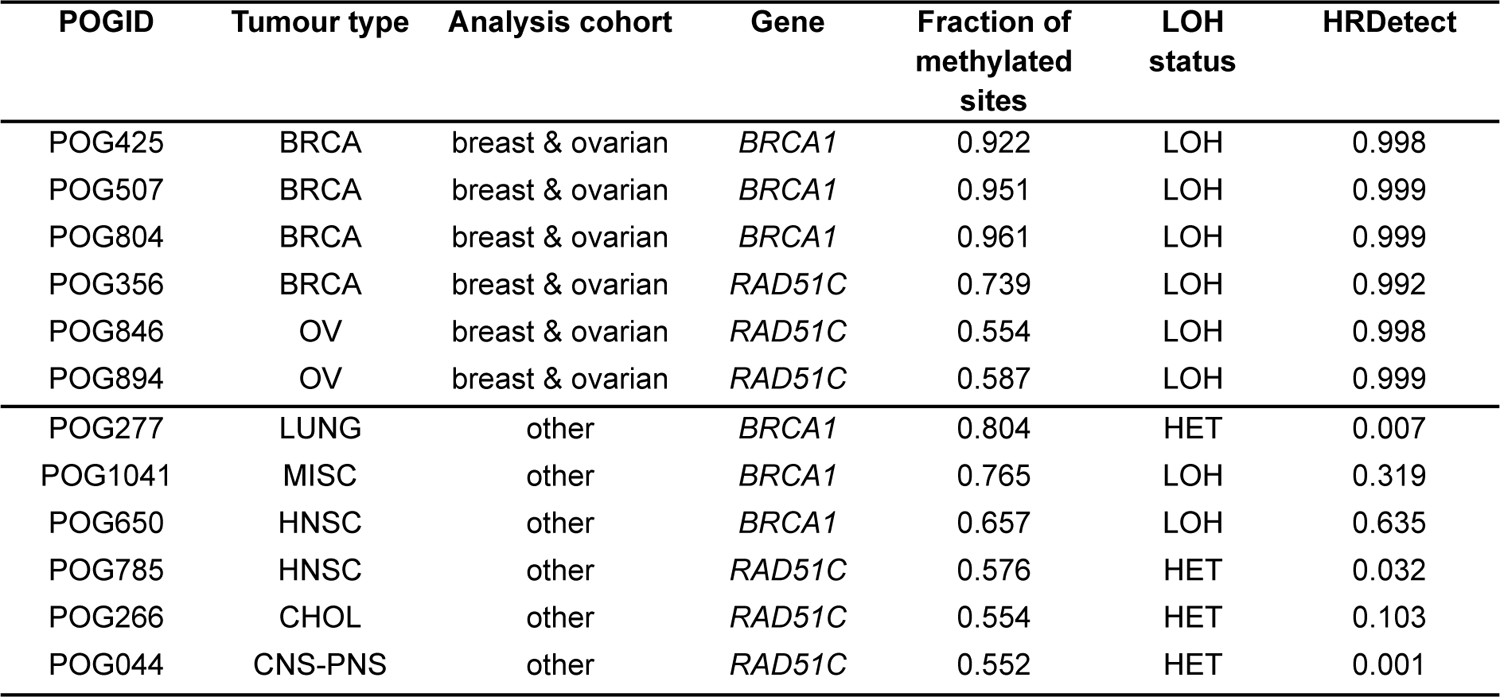
Promoter methylation of HR genes in cases with HRD.

Compared to the rest of the cohort, samples with *BRCA1* or *RAD51C* promoter hypermethylation showed reduced gene expression. This is consistent with methylation-induced transcriptional silencing (Figure 5b). Haplotype-specific methylation data revealed that the methylation was confined to a single allele in the HRD tumours with *BRCA1* (Figure 5c) and *RAD51C* (Figure 5d) promoter hypermethylation. Together with the observed loss of heterozygosity of the other allele, this is consistent with biallelic inactivation of these genes. Matched blood was available for all three *BRCA1* and one of the *RAD51C* cases. In all instances the promoter alleles were unmethylated in the blood, suggesting that the tumour promoter methylation was a somatic event (Figure 5c-d).

HRD is observed in other cancers, albeit at lower frequencies. However, *BRCA1* and *RAD51C* promoter hypermethylation have only been reported in sporadic breast and ovarian cancer cases^50,51^. In this cohort we identified promoter hypermethylation of *BRCA1* and *RAD51C* in a further three cases each (Table 2). Two of the samples with *BRCA1* promoter hypermethylation showed LOH of *BRCA1* and exhibited moderate HRDetect scores, suggesting potential HRD. In the remaining four samples, the other allele was intact and unmethylated, and the HRDetect score was low (Table 2), likely due to incomplete inactivation of the gene (Extended Data Figure 9a-c).

Outside of Lynch syndrome, inactivation of mismatch repair (MMR) genes and consequent microsatellite instability arises in sporadic cancers, usually from a combination of somatic MMR gene mutations or somatic hypermethylation at the promoter region. However, constitutional epimutations that result in germline promoter hypermethylation have been reported^52^. Our cohort included a patient, POG986, with lung squamous cell carcinoma and with multiple other previous primary cancer diagnoses, suggestive of Lynch syndrome. Previous clinical hereditary cancer multigene panel sequencing was uninformative, but targeted methylation testing of blood performed after short-read POG analysis showed constitutional methylation of MLH1 confirming Lynch syndrome. Long-read sequencing data from this study further confirmed monoallelic hypermethylation in the blood with no causative sequence variants (Figure 5e). In the tumour, somatic LOH of *MLH1* resulted in the loss of the wild-type *MLH1* allele, with hypermethylation on the remaining tumour allele (Figure 5e,f, POG986). Another tumour, endometrial cancer POG041, showed hypermethylation on both tumour alleles of *MLH1* (Figure 5f). MMR deficiency was confirmed by immunohistochemical testing. No matched blood sample was long-read sequenced, but the absence of a germline mutation and the presence of biallelic methylation in the tumour suggest that the *MLH1* promoter methylation was a somatic event in this case.

### Genomic and epigenomic architecture of extrachromosomal DNA

We hypothesized that the added methylation and phasing information obtained from long reads would enable the reconstruction of both the genomic and epigenomic structure of extrachromosomal DNA (ecDNA). Using AmpliconArchitect^53^ on the short-read WGS data, we predicted the presence of 76 ecDNAs in 42/189 (22.2%) samples (Supplementary Table 1). A total of 1,283 genes were detected on the ecDNAs in our cohort, with 262 (20.4%) occurring on more than one ecDNA (Supplementary Table 6). Importantly, 97 of these genes were oncogenes, 33 (34.0%) of which were recurrent in our cohort. *ZNF703* recurred most frequently, being present in ecDNAs in five samples. The presence of at least one ecDNA correlated with an increased genomic complexity score (two-sided Student’s *t*-test; adjusted *P*=5.17×10^-7^) and a trend toward an increased HRD score (two-sided Student’s *t*-test; adjusted *P*=0.0548), which may be partially explained by the increased representation of breast cancers in the ecDNA+ cohort (Extended Data Figure 10b).

We examined allelic methylation patterns within promoter regions for genes predicted to reside on ecDNAs, as a potential mechanism of ecDNA-specific dysregulation. We identified one breast cancer case with multiple aDMRs within an ∼4 Mb ecDNA, surrounding *NRG1.* Two aDMRs overlapped promoter regions for separate isoforms of the gene (Figure 5g). *NRG1* is a known cancer gene in breast cancer^54–57^. We were able to validate 4/5 of the breakpoints predicted by AmpliconArchitect by manual review of the long-read data (see Methods). We were further able to assign the ecDNA to a single haplotype based on the nanopore long-range phasing, and to confirm that the entire event fell within one phase block (Figure 5h; see Methods). Both aDMRs in *NRG1* promoter regions showed hypermethylation of the amplified (ecDNA) allele. Furthermore, across the entire ecDNA region, the density of aDMRs was significantly greater within *NRG1* than any other region of the ecDNA (Figure 5i). Although *NRG1* was expressed only from HP1, and was focally amplified in the genome, it was not overexpressed relative to other breast cancer samples within the cohort (Extended Data Figure 10d). This finding suggests a regulatory mechanism by which an ecDNA-mediated amplification may be countered by promoter methylation of the amplified gene.

## Discussion

We present the Long-read POG Cohort, a set of 189 tumour samples of diverse tumour types sequenced via the Oxford Nanopore Technologies sequencing platform. As the samples in this cohort have accompanying short-read DNA and RNA sequence data and associated clinical information, our study offers potential for advancing the understanding of SVs, viral integration, DNA methylation, and allelic information pertinent to cancer pathogenesis and diagnosis.

Long-read sequencing data enabled automatic calling of known oncogenic fusions, and showed reasonable concordance with short-read SV calls genome-wide. When examined, discordance was largely due to the much lower coverage of long reads, or inability to map breakpoints in the short reads. Long-read data enabled reconstruction of complex SV events that were undetected or mischaracterized in short-read data. Furthermore, long-read data enabled the direct detection of MSI expansions as SVs^22,23^, as well as the detection of “tyfona” signatures in sarcoma and melanoma as previously described^24^. Complex patterns of HPV integration could also be deconvoluted, along with their impact on surrounding cancer gene expression, as has been recently explored in cervical cancer^58,59^. Currently, there is limited availability of somatic SV callers for the nanopore platform, but with the aid of datasets such as the Long-read POG cohort, development of additional calling approaches will allow for improvement in this area.

Long-range phasing of variants using long reads facilitates the coverage of the vast majority of genes within single haplotype blocks. This enables phasing of double hits in tumour suppressors to ascertain biallelic loss of function. Moreover, when combined with short-read RNA data, phasing can link deleterious variants with ASE, providing further confirmation of multiple hits to tumour suppressors. We observed widespread recurrent ASE across tumours, as exemplified by the widespread ASE of *DUSP22*. The re-expression in tumours of genes which are typically developmentally restricted, such as *DUSP22*, has been suggested to be a potential source of neoantigen therapeutic targets^60^.

The ability of nanopore sequencing to provide methylation data within standard WGS without additional sequencing or bisulfite conversion is a significant advantage. We showed that methylation is associated with tissue of origin, suggesting the potential for tumour type classification^61^. Long-range phasing of methylation facilitates the exploration of aDMRs. Although we confirmed that the majority of aDMRs are driven by CNVs, we also observed epigenetic dysregulation in copy-neutral regions. In rare cases, this can be due to germline altered methylation, exemplified by the identification of germline inactivating *MLH1* methylation in a patient with clinically confirmed Lynch syndrome. This phenomenon has been described in both familial and sporadic Lynch syndrome patients, along with acquired hypermethylation of *MLH1* in endometrial cancers^62^. Long-read sequencing could be applied to identify causative epigenomic alterations in Lynch and other syndromes.

We observed recurrent aDMRs in the intragenic promoters of *RET* and *CDKN2A,* with methylation being associated with increased expression of the canonical transcript. The effect of intragenic promoter methylation on transcription is complex and bidirectional^63–65^. Gene bodies frequently become methylated during active transcription^63,64^, which may silence intragenic promoters via transcription interference^63–65^. Conversely, intragenic promoter methylation can increase canonical transcript expression by reducing competition for RNA Pol binding, or through regulating transcription elongation^63^, with evidence that this methylation regulates some oncogenes in cancer^66,67^. Further analysis of intragenic promoter methylation in *RET* and *CDKN2A* is needed to determine whether this change is a mere consequence of active transcription or a key regulator of expression. Promoter methylation in the oncogene *NRG1* in one case was also notable for being ecDNA-specific, a phenomenon which has not been well characterised to date^73^.

Our study expounds the significant role of gene promoter hypermethylation in HRD tumours, which often lack a clear underlying causative mutation^68,69^. Notably, the presence of *BRCA1* and *RAD51C* promoter methylation was associated with a clinically-relevant HRD phenotype, when accompanied by a second hit, typically by LOH. Interestingly, *BRCA1* promoter hypermethylation was observed in melanoma and head & neck cancer, in addition to breast and ovarian cancers where it has been typically described^50,51^. This suggests that HRD gene promoter methylation may be involved in a broader spectrum of tumours, a clinically important finding as deficiency in HR is associated with sensitivity to platinum chemotherapies and PARP inhibitor therapies^70,71^.

In conclusion, we present a comprehensive cohort of tumours sequenced on the nanopore platform. Our initial findings suggest a role for long-read sequencing in personalized cancer medicine through the phasing of somatic mutations, deconvolution of structural variation, identification of tumours driven by HRD, and the discovery of allele-specific methylation of tumour suppressors. The single long-read approach has advantages for clinical use including providing both sequence and methylation information with comparatively simple library preparation and rapid turnaround times^74^. Additional tumour long-read sequencing is warranted as a complement to the established short-read sequencing paradigm to understand its use in biomarker-driven clinical trials and identifying targeted treatment options. We provide this dataset, complemented by clinical information and short-read sequencing data, as a valuable resource for benchmarking, annotation, and fostering continuous improvement in cancer research and clinical practice.

## Data Availability

Genomic and transcriptomic sequence datasets for long-read and short-read platforms have been deposited at the European Genome-phenome Archive (EGA, https://ega-archive.org/) as part of the study EGAS00001001159 with accession numbers as listed in Supplementary Table 1. Processed data from Long-read POG, figure source data and accompanying short-read variants can be downloaded from https://www.bcgsc.ca/downloads/nanopore_pog/. Data on mutations, copy changes and expression from tumour samples in the POG program are also accessible from https://www.personalizedoncogenomics.org/cbioportal/. Code used to generate figures in this manuscript is available in containerized, reproducible form at https://github.com/bcgsc/long_read_pog. WGBS data, ENCODE accessions and samples from GSE186458 that were used as normal tissues are included in Supplementary Table 7.

## Acknowledgments

This work would not be possible without the participation of our patients and families, the POG team, Canada’s Michael Smith Genome Sciences Centre technical platforms, the generous support of the BC Cancer Foundation and their donors, the Terry Fox Research Institute Marathon of Hope Cancer Centres Network, and Genome British Columbia (project B20POG). We acknowledge contributions from Genome Canada and Genome BC (projects 202SEQ M.A.M & S.M.J, 212SEQ M.A.M & S.M.J, 12002 GBC M.A.M, S.M.J & J.L), Canada Foundation for Innovation (projects 20070 M.A.M & S.M.J, 30981 M.A.M, S.M.J & J.L, 30198 M.A.M, 33408 M.A.M & S.M.J, 40104 M.A.M & S.M.J, 42362 S.M.J) including the CGEN platform (35444 S.M.J) and the BC Knowledge Development Fund. We acknowledge the generous support of the CIHR Foundation Grants program (FDN 143288, M.A.M). The results published here are in part based upon analyses of data generated by the following projects and obtained from dbGaP (http://www.ncbi.nlm.nih.gov/gap): Genotype-Tissue Expression (GTEx) Project, supported by the Common Fund of the Office of the Director of the National Institutes of Health (https://commonfund.nih.gov/GTEx).

## Author Contributions

**Wrote the manuscript:** Kieran O’Neill, Erin Pleasance, Jeremy Fan, Vahid Akbari, Glenn Chang, Katherine Dixon, Veronika Csizmok, Signe MacLennan, Vanessa Porter, Andrew Galbraith

**Carried out analyses and prepared figures:** Kieran O’Neill, Erin Pleasance, Jeremy Fan, Vahid Akbari, Glenn Chang, Katherine Dixon, Veronika Csizmok, Signe MacLennan, Vanessa Porter, Andrew Galbraith, John Dupuis

**Carried out broader/supporting analyses:** Richard Corbett, James Hopkins, Reanne Bowlby, Dean Cheng, Tina Wong, Connor Frey, Yaoqing Shen, Luis F Paulin

**Developed, coordinated and carried out custom lab work:** Pawan Pandoh, Duane Smailus

**Supervised the work:** Kieran O’Neill, Erin Pleasance, Fritz Sedlazeck, Jessica M.T. Nelson, Eric Chuah, Karen L. Mungall, Richard A. Moore, Robin Coope, Andrew J. Mungall, Melissa K. McConechy, Laura M. Williamson, Kasmintan Schrader, Stephen Yip, Marco A. Marra, Janessa Laskin, Steven J.M. Jones

## Competing interests

The following authors disclose relevant potential competing interests: Kieran O’Neill, Vanessa Porter, Luis F Paulin, Katherine Dixon and Janessa Laskin and Steven J.M. Jones received travel funding from Oxford Nanopore Technologies to present at conferences in 2022 and 2023.

## Methods

### Enrollment and clinical data collection

The POG program, registered under clinical trial number NCT02155621, was approved by the University of British Columbia – BC Cancer Research Ethics Board (H12-00137, H14-00681), and approved by the institutional review board. As described in Pleasance et al^6^, patients were referred by their treating oncologist in British Columbia, Canada, and enrolled based on criteria including locally advanced or metastatic cancer, predominantly having received one or more lines of therapy in the metastatic setting, ECOG ≤ 1, life expectancy >6 months, and the ability to undergo biopsy procedures. POG IDs (POGXXXX) were assigned to each case as anonymous identifiers known only to the research group. Samples with availability of sufficient nucleic acid material after short-read sequencing were considered for nanopore sequencing.

Overall survival was evaluated from the date of advanced disease diagnosis, defined as the date of incurable, advanced or metastatic disease as determined by radiology or clinical finding if progression was documented with subsequent imaging, whichever was earlier. Kaplan-Meier survival analysis was performed as of August 1, 2023 using the R packages survival (v2.42.3) and survminer (v0.4.2).

### Sample preparation and sequencing

#### Extraction and size selection

Nucleic acids for this study were obtained from previously purified samples from tissues accompanied by tumour estimates as described in Pleasance et al^2^. Purification was performed with an A-Line Evo-pure kit automated on a Hamilton Nimbus96 robot. The overall workflow and automated steps are shown in Figure 1C and Extended Data Figure 1b. Briefly, frozen tissue sections were immersed in 420 μL of RLT Plus buffer (QIAGEN) containing tris(2-carboxyethyl)phosphine (a reducing agent; TCEP) and a unique sample tracking DNA plasmid, and gently agitated overnight at room temperature. Lysates were transferred from 2 mL tubes to wells of a 1.2 mL plate (Thermo Scientific, AB1127) to which was added 400 mL of 5x bind buffer (80 mL beads in 320 mL isopropyl alcohol). Following a 5 minute incubation at room temperature lysates were cleared on a Magnum FLX magnet place (Alpaqua Inc) for 6 minutes and the protein-containing supernatant removed. The beads, with bound nucleic acids, were washed by pipetting 10 times in wash buffer and returned to the magnet. Beads were washed three times in 70% ethanol then dried for 10 minutes. 40 mL nuclease-free water was added to the dried beads and returned to the magnet. The eluted total nucleic acids were transferred to a 96-well storage plate and aliquots taken for quantification using Invitrogen Qubit 4 Fluorometer (Thermo Fisher Q33226)

For samples with concentrations below 166 ng/µL or in volumes greater than 30 µL, 5000 ng of Total Nucleic Acid (TNA) was transferred to a 1.5 mL DNase/RNase free tube and concentrated without heat on the Savant SpeedVac Plus (SC210A) to a maximum volume of 30 µL. This 5000 ng sample in 30 µL was topped up with 10 µL of Qiagen Buffer EB (Cat.19086). The samples were then run on the SAGE Science Blue Pippin instrument using the High Pass Plus Cassette (BPPLUS 10) with a maximum of 4 samples per run. The start range was selected at 15,000 base pairs and the end range was selected at 150,000 base pairs yielding a targeted size of 82,500 base pairs. Following the run completion, each sample was eluted in 80 µL and each sample elution well was then washed with 80 µL of 10 mM Tris, 1mM EDTA containing 0.1% Tween to maximize sample recovery. The total volume of 160µL was then concentrated without heat to 50 µL using the SpeedVac Plus. Each sample was then quantified using the Invitrogen Qubit 4 Fluorometer (Q33226) and normalized to 2000 ng in 47.5 µL for PromethION genome sequencing.

#### Library construction and sequencing

Library construction and sequencing followed the Oxford Nanopore Technologies Genomic DNA by Ligation (SQK-LSK110) protocol. DNA libraries starting with 2 µg of sample per library. No shearing was performed. The NEBUltra-II kit, (New England Biolabs, Ipswich, MA, USA, cat. no. E7646A) was used for end-repair and A-tailing. NEBNext quick ligase (E6056S) was used to ligate the Oxford Nanopore sequencing adapter. A final size selection of 0.4:1 ratio beads:library (PCRClean – DX Aline Biosciences L/N 06180316) was done to select against smaller molecules. These library preparation steps were performed on Hamilton Nimbus96 liquid handlers. An example deck layout, in this case for the bead purification step, is shown in Extended Data Figure 1c. DNA libraries were loaded in R9.4.1 pore flow cells on PromethION 24 instrument running software version 19.06.9 (MinKNOW GUI v4.0.23). Sequencing was carried out for 72 hours. DNase I (Invitrogen cat no. AM2222) nuclease flush was performed after 24-48 hours by reloading the flow cell with the same library mix.

#### Basecalling and Primary Analysis

Basecalling was performed using the guppy basecaller from Oxford Nanopore Technologies, using the “super-accurate” model. Primary analysis was carried out using a NextFlow workflow, which is provided at https://github.com/bcgsc/long_read_pog. Small variants were called using clair3^75^ (v0.1-r8) and phased using WhatsHap as included with clair3. Structural variants were called using sniffles^76,77^ (1.0.12b) and cuteSV^78^ (1.0.12). Methylation (5-mC) was called using nanopolish^13^ (0.13.3) and phased using nanomethphase^79^.

### Short-read data analysis

Short-read data from the Illumina platform was generated as described in Pleasance et al^2^. Reads were aligned to the human reference genome (hg38) using Minimap2^80^ (v2.15). Regions of copy number variation and losses of heterozygosity were identified using Ploidetect (https://www.biorxiv.org/content/10.1101/2021.08.06.455329v1) (v1.3.0 and v1.4.2). Tumour content (estimated proportion of DNA derived from tumour cells vs normal cells in the sample) and average ploidy observed in the sequenced tumour were determined based on manual review of Ploidetect results, copy number plots and allelic ratios. Two measures of copy number complexity were computed: the fraction of the genome falling in non-ploidy copy segments, and the genome complexity which is the arithmetic mean of the fraction non-ploidy and the fraction of the total genome size falling in non-ploidy segments, computed based on Ploidetect copy number results with segments less than 10kb merged. Somatic single nucleotide variants (SNVs) and small insertions and deletions (indels) were identified using Strelka2^81^ (v2.9.10) and Mutect2 (https://www.biorxiv.org/content/10.1101/861054v1) (in GATK v4.2.0.0). Events assigned PASS by both callers were included, as well as indels called by Strelka2 only with QSS>=50. Tumour mutation burden (TMB) was computed as total called somatic SNVs and indels per megabase. Somatic structural variants (SVs) in DNA data were identified using assembly-based tools ABySS (v1.3.4)^82^ and Trans ABySS (v1.4.10)^82,83^ and alignment-based tools Manta (v1.6.0)^84^ and Delly (v0.8.7)^85^, with consensus calls merged using MAVIS^86^ (v2.2.1). SVs were filtered to exclude those with identical genomic breakpoints in multiple samples, to remove from the somatic call set germline variants and some technical artifacts. SV events were considered high quality (HQ) if they were called by more than one tool and if a contig could be assembled that aligned across the candidate genomic breakpoint, otherwise they were classed as low quality (LQ). Variants were annotated to genes using SNPEff^87^ (v5.0) with the Ensembl database^88^ (v100). MSI samples were identified with MSIsensor^89^ (v2.0.1). Microbial detection was performed using BioBloomTools (v2.0.11b)^21^. RNA-Seq reads were aligned using STAR^90^ (v2.5.2b-XS and Sambamba 0.7.1) and expression was quantified using RSEM^91^ (v1.3.0) based on gene models from Ensembl v100.

### Structural variation characterization

#### SV identification and processing

We conducted two distinct analyses. We performed tumour-only SV calling for all tumours (*n*=189). For the subset of tumours with matched normal, (*n*=43), we performed somatic SV calling. A literature review was conducted as of May 2023 of existing long reads somatic SV callers, and callers were selected based on the criteria that they had detailed documentation and were continually being maintained over the last year. We identified two somatic SV callers meeting these criteria: SAVANA(v1.0.3) (https://github.com/cortes-ciriano-lab/savana) and nanomonSV(v0.5.0)^20^. CuteSV(v1.0.12)^78^ and Sniffles(v2.0.7)^76,77^ were used as germline SV callers for the tumour-only analysis. Callers were run with default parameters and a minimum size threshold of 50 bp. Intrachromosomal breakend notation for SAVANA calls were transformed to different SV type calls according to VCF4.2 conventions. Passing SAVANA and nanomonSV events fulfilling a minimum tumour variant allele frequency (VAF) of 0.05 with no event support in the matched normal were subsequently used for downstream analysis.

Post-processing of SVs was conducted with MAVIS (v.3.1.0)^86^. We filtered SVs found in the sex and unknown chromosomes, collapsed duplicate SVs, and merged/clustered SVs by breakpoint proximity (100 bp) and type. Insertion end coordinates were calculated by adding the length of the inserted sequence alongside the confidence interval range. SVs called somatic but appearing in paired normals were categorised as false positives and filtered out. SVs were annotated using events from the Database of Genomic Variants (DGV)^92^ and frequent events seen in normal WGS using MAVIS. SVs were also annotated based on RepeatMasker (v4.1)^93^.

#### SV comparisons

Events were considered to match known fusions if the SV had both breakpoints within 10 bases upstream or downstream of the reported breakpoints and were classified as the same SV type. Events selected for manual review were those that overlap a gene found in OncoKB^94^ and which did not have breakpoints overlapping within a repetitive element in RepeatMasker. To visualise somatic events in regions of interest, we took the SAVANA SV calls alongside the Ploidetect(v1.3.0) copy number calls into ShatterSeek^95^.

Corresponding short-read somatic MAVIS post-processed SVs from BreakDancer, DeFuse, Manta, Delly, Trans-ABySS, and ChimeraScan were compared to nanopore SVs. Somatic cohort wide level calls were analysed for any SVs spanning coding elements of oncogenic genes within OncoKB. SVs were considered similar if they clustered within 100 bp of each other. To resolve complex events, we pulled out all reads surrounding the region of the hypothesised event, and took the majority of the reads which supported a certain interpretation. Afterwards, we conducted a local assembly of the reads involved in the structural variants (determined by all reads that support an event in the proximal distance of the complex event. Finally, we assessed whether the regions by subsetting for HQ Illumina events and nanopore events, look for those which predict a non-synonymous coding change from short reads and those events which overlap known tumour suppressors or oncogenes. We manually reviewed the transcriptomic data for evidence of irregular splicing patterns and expression profiles through sashimi plots.

### Viral Integration

HPV viral breakpoints were detected as described by Porter et al^96^ and using the workflow from GitHub (https://github.com/vanessa-porter/callONTIntegration). Briefly, Sniffles (1.0.12) was used to call breakpoints as translocations between the human chromosomes and the HPV genomes using a minimum of 5 consensus reads. Breakpoints were grouped together into HPV integration events if they had one or more shared reads or mapped within 500 kb of each other as measured using BEDTools (v. 2.30.0). Integration event structures were determined using the read alignment patterns as described by Porter et al, 2023 (https://doi.org/10.1101/2023.11.04.564800). The multi-breakpoint event was analysed using the workflow found here: https://github.com/vanessa-porter/comSVis, which sectioned the event using the collection of all SV breakpoints that were phased within the event. The mean depth between the breakpoints was then calculated using BEDTools (v. 2.30.0) for visualisation.

### Phasing

Individual phasing statistics were calculated for each phased VCF using WhatsHap stats. Read N50 is the length at which reads of the same or greater length represent 50% of the genome. To estimate the phasing rate across tumour suppressor genes, we determined the number of protein-coding genes (GENCODE v43) that are contained within a single phase block for each sample using bedtools intersect, restricting overlapping genes to those that had a 100% overlap with a given phase block. Putative biallelic somatic variants with potential biological or clinical significance were identified from the POG genomic reports.

Tumour suppressor genes were defined by the COSMIC Cancer Gene Census. Long reads were coloured by predicted haplotype using WhatsHap haplotag, and all candidate biallelic variants were manually reviewed in IGV.

### Allele Specific Expression

The IMPALA pipeline (github.com/bcgsc/IMPALA; https://www.biorxiv.org/content/10.1101/2023.09.11.555771v1) was used to detect ASE genes in the POG cohort (*n*=172), which uses tumour RNA-seq data and phased variants generated from tumour long reads. STAR aligner^90^ (v2.7) is used to align the RNA reads to the genome before performing variant calling using Strelka (v2.9). This generates allelic read counts for each variant. Heterozygous SNPs are filtered for and annotated with haplotype information and gene annotation using the phased variants and SnpEff^87^ (v5.0) respectively. SnpSift (v5.1d) formats the allelic read count and annotations as preprocessing for ASE detection. RSEM^91^ (v1.3) is used to quantify expression of RNA-seq data and filter out genes with expression lower than 1 TPM.

MBASED^97^ (v1.34) is the main software used to calculate ASE. Biallelic genes are expected to have an allelic read count ratio of 0.5. MBASED performs a beta-binomial test on each phased SNP to assess the statistical deviation away from the expected 0.5 ratio. Afterwards, MBASED utilises meta-analysis with haplotype information to aggregate SNP-level data to gene-level major allele frequency data. P-value is adjusted with Benjamini-Hochberg method. Genes with major allele frequency above 0.65 and adjusted p-value below 0.05 are classified as allelically expressed.

Post-processing of the allele specific expression is done by integrating additional information to determine the potential cause of the allele specific expression. CNV data from Ploidetect, allelic methylation data from NanoMethPhase and somatic variant calls can be used as optional inputs for IMPALA software. Bedtools (v2.23) intersect is used to intersect ASE data with CNV states, allelic methylation and somatic calls. Additionally, SnpEff is used to annotate and filter for nonsense variants in ASE gene as a potential genetic mechanism. Lastly, bcftools (v1.15) consensus is used to generated consensus sequence of both allele based on the phased variants and FIMO from the MEME suite (v5.4.1)^98^ detects transcription factor binding sites on both alleles and find differences between the allele. Disruption of transcription factor binding sites could lead to ASE. The final output of the workflow is a summary table with allelic information in addition to the cis-acting elements which can be used for downstream analysis to identify genes of interest.

### Methylation analysis

For non-allelic methylation analysis we used nanopolish methylation frequency results. As normal methylation data, 267 WGBS datasets from various tissue/cell types were gathered from Epigenomics Roadmap^99^, ENCODE^100^, and Loyfer et al^101^ (GSE186458). To analyze overall methylation at different genomic regions, we used bedtools intersect to overlap CpG methylation frequency data to Repeats, TF binding sites (also includes CTCF binding sites), and CGIs from the UCSC table browser^102^, promoters (1500 bp upstream and 500 bp downstream of TSS in Ensembl100 transcripts GRCh38.p13^88^), 500bp up and downstream of polyA sites from PolyA_DB 3^103^, and enhancers from GeneHancer v5.14^104^. For tSNE, we used CGIs, Promoters, CTCF binding sites, and Enhancers regions with standard deviation≥0.2 of the mean methylation between tumour types or biopsy sites. For both phased and unphased data, if the methylation was at strand level, for each CpG site we aggregated the number of reads as methylated and number of all reads from both strands to calculate consensus methylation frequency.

To detect aDMRs in each sample, the phased haplotype 1 and 2 results from NanoMethPhase were used and DMRs were called in each sample using NanoMethPhase dma module with default options with DSS R package version 2.46.0^79,105^. Detected DMRs were further filtered to keep DMRs with |diff.methyl (delta methylation)| ≥0.15. To filter detected aDMRs in tumour samples and keep only tumour-specific aDMRs, in addition to ignoring aDMRs that overlap to more than one matched blood sample, we excluded aDMRs showing partial methylation in more than 1% of the normal WGBS samples. Partial methylation is an indication of allelic methylation because only one allele is methylated and overall methylation at the region will be ∼50%. To explore partial methylation, for each WGBS sample, we used CpGs with at least five mapped reads and at each detected aDMR we counted the number of CpGs with partial methylation (methylation frequency between 0.35 and 0.65). An aDMR with 0.35–0.65 methylation is then considered partially methylated if it had at least five CpGs in the WGBS sample and more than 60% of the CpGs showed partial methylation. To overlap detected aDMRs to genomic regions we used bedtools intersect -e -f 0.5 -F 0.5. TF enrichment for recurrent genes with aDMR at their promoter was evaluated using the Enrichr ChEA 2022 database^106^ (https://maayanlab.cloud/Enrichr/). CIBERSORT version 1.6.2 was used to infer immune infiltrate proportions using gene expression data.

### HRDetect and HR gene promoter methylation

We used HRDetect, a tool which aggregates different mutational signatures including single base substitution signatures, structural variant signatures and microhomology-mediated deletions, to predict HRD in our samples. HRDetect scores were computed from short-read sequencing data using a logistic regression model with the same intercept and coefficients as those reported in the previously trained model, without adjustment^107^. The intercept was −3.364 and the coefficients were 1.611, 0.091, 1.153, 0.847, 0.667, and 2.398, respectively, for the six HRD signatures: (i) SBS3, (ii) SBS8, (iii) SV signature 3, (iv) SV signature 5, (v) the HRD index, and (vi) the fraction of deletions with microhomology. The contribution of previously reported mutational signatures in the Catalogue of Somatic Mutations in Cancer (COSMIC v3.1, https://cancer.sanger.ac.uk/cosmic/signatures) was calculated using Monte Carlo Markov Chain (MCMC) sampling (https://github.com/eyzhao/SignIT). Short-read MAVIS calls that were detected by more than one tool and for which the contig could be assembled were included in the analysis and the contribution of the previously reported SV mutational signatures was calculated using MCMC sampling (https://github.com/eyzhao/SignIT)^108^. The HRD index was computed as the arithmetic sum of loss of heterozygosity, telomeric allelic imbalance, and large-scale state transitions scores. The microhomology fraction was determined as the proportion of deletions which were larger than three base pairs and demonstrated overlapping microhomology at the breakpoints^70^. All signatures were log transformed and normalized so that each feature had a mean of 0 and standard deviation of 1^107^.

Promoter methylation of the following HR genes, selected based on their established roles in homologous recombination repair, were investigated to examine associations with high HRDetect score: *BARD1, BLM, BRCA1, BRCA2, BRIP1, DNA2, EXO1, MRE11A, NBN, PALB2, RAD50, RAD51, RAD51B, RAD51C, RAD51D, RAD52, RAD54L, RBBP8, WRN, XRCC2, XRCC3, ATM, BAP1, CUL3, EME1, ERCC1, ERCC4, FBXO18, GEN1, HELQ, MUS81, PARPBP, PCNA, POLD1, POLK, POLN, PSIP1, RAD51AP1, RECQL5, RIF1, RMI1, RMI2, RPA1, RPA2, RPA3, RTEL1, SLX1A, SLX4, TOP3A, TP53BP1,* and *USP11*. Promoter is defined as 1500 bp upstream and 500 bp downstream from TSS. Methylation frequencies from Nanopolish in the promoter regions of 51 HR genes in the tumours and in publicly available matched tissues (https://epigenomesportal.ca/ihec/) were compared. ‘Methylated’ site in the promoter is defined as that methylation of the site being >1 SD from the mean normal methylation level in the matched tissue and then the fraction of methylated sites in the gene promoter is computed.

### Extrachromosomal DNA characterization

#### ecDNA detection and visualization

To identify potential ecDNAs from short-read WGS data, we ran PrepareAA (v0.1203.1) with CNVkit^109^ (v0.9.10.dev0) for copy number calling followed by AmpliconArchitect^53^(v1.2) with default settings (--gain 4.5 and --cnsize_min 50000). For ecDNA structure visualization, we first used CycleViz (v0.1.2; github.com/AmpliconSuite/CycleViz) to obtain breakpoints predicted by AmpliconArchitect for ecDNAs with only one predicted substructure, followed by circos^110^ (v0.69.9), in which we overlaid additional methylation data obtained from long-read WGS data.

#### ecDNAs and survival

For the 181 patients in the cohort, we used diagnosis and, if applicable, death dates to construct Kaplan-Meier survival curves stratified on ecDNA presence using the survival^111^ (v3.5-5) and ggsurvfit^112^ (v0.1.0) packages in R (v4.0.2) with lubridate^113^ (v1.9.2) and tidyverse^114^ (v2.0.0) for data preparation. A log-rank test was used to assess significance between ecDNA+ and ecDNA-survival curves. A Fisher’s exact test was also used to compare the number of surviving vs dead patients with cancers containing ecDNAs vs those without ecDNAs.

#### Validation of ecDNA structure using long reads

To validate the structure of select ecDNAs predicted by AmpliconArchitect^53^, which uses short-read WGS data, we manually reviewed supplementary reads from the long-read WGS data in IGV^115^ (v2.14.1). Specifically, we looked for reads mapping to both sides of each predicted breakpoint +/- 100 bp. We assigned an ecDNA to a specific haplotype based on whether reads mapping to SVs associated with the ecDNA mapped to reads within the haplotype-phased bam file. Specifically, we extracted SVs associated with the ecDNA from the output of AmpliconArchitect, found these SVs in the long-read WGS data from Sniffles (v1.0.12b), and then mapped these reads to both tumour haplotype bam files. For further validation of the assignment of an ecDNA to a specific haplotype, we viewed the ecDNA in IGV to confirm amplified regions co-localized with the ecDNA regions of AmpliconArchitect.

#### Overlapping DMRs with ecDNAs

We used annotatr^116^ (v1.16.0) in R (v4.0.2) to extract gene promoters overlapping both ecDNA regions and DMRs obtained from the allele-specific methylation pipeline prior to filtering out CNVs (see also Allele-specific methylation). We selected *NRG1* in a breast cancer sample, for further analysis as it is a known cancer gene in breast cancer^54–57^, had multiple DMRs falling within it, including two in promoter regions, and had >0.5 methylation frequency for one haplotype and < 0.5 methylation frequency for the other haplotype for each promoter DMR. Plots for *NRG1* methylation were constructed in R with tidyverse^114^ (v2.0.0) and patchwork (v1.1.2.9000)^117^ functions with ggbio^118^ (v1.38.0) and EnsDB.Hsapiens.v86 (v2.99.0) for gene annotation.

#### NRG1 expression analysis

We compared *NRG1* and other genes within the *NRG1* pathway (*ERBB2*, *ERBB3*, and *AKT1*) between the sample containing the *NRG1* ecDNA (*n*=1) to other breast cancers within the cohort (*n*=39) in terms of RNA expression in TPM. Permutation tests were used to assess significance between the ecDNA sample and the rest of the breast cancers in the cohort using the coin^119^ (v1.4-2) package and Bonferroni multiple testing correction. We also reviewed ASE results for *NRG1* in the sample of interest (see also Allele-specific expression).

## Notes

### Competing Interest Statement

The following authors disclose relevant potential competing interests: Kieran ONeill, Vanessa Porter, Luis F Paulin, Katherine Dixon and Janessa Laskin and Steven J.M. Jones received travel funding from Oxford Nanopore Technologies to present at conferences in 2022 and 2023.

### Funding Statement

This work would not be possible without the participation of our patients and families, the POG team, Canadas Michael Smith Genome Sciences Centre technical platforms, the generous support of the BC Cancer Foundation and their donors, the Terry Fox Research Institute Marathon of Hope Cancer Centres Network, and Genome British Columbia (project B20POG). We acknowledge contributions from Genome Canada and Genome BC (projects 202SEQ M.A.M & S.M.J, 212SEQ M.A.M & S.M.J, 12002 GBC M.A.M, S.M.J & J.L), Canada Foundation for Innovation (projects 20070 M.A.M & S.M.J, 30981 M.A.M, S.M.J & J.L, 30198 M.A.M, 33408 M.A.M & S.M.J, 40104 M.A.M & S.M.J, 42362 S.M.J) including the CGEN platform (35444 S.M.J) and the BC Knowledge Development Fund. We acknowledge the generous support of the CIHR Foundation Grants program (FDN 143288, M.A.M). The results published here are in part based upon analyses of data generated by the following projects and obtained from dbGaP (http://www.ncbi.nlm.nih.gov/gap): Genotype-Tissue Expression (GTEx) Project, supported by the Common Fund of the Office of the Director of the National Institutes of Health (https://commonfund.nih.gov/GTEx).

### Author Declarations

The POG program, registered under clinical trial number NCT02155621, was approved by the University of British Columbia - BC Cancer Research Ethics Board (H12-00137, H14-00681), and approved by the institutional review board.

## References

1. Priestley, P. et al. Pan-cancer whole-genome analyses of metastatic solid tumours. Nature 575, 210–216 (2019).

2. Pleasance, E. et al. Pan-cancer analysis of advanced patient tumors reveals interactions between therapy and genomic landscapes. *Nat*. Cancer 1, 452–468 (2020).

3. Zehir, A. et al. Mutational landscape of metastatic cancer revealed from prospective clinical sequencing of 10,000 patients. Nat. Med. 23, 703–713 (2017).

4. Chalmers, Z. R. et al. Analysis of 100,000 human cancer genomes reveals the landscape of tumor mutational burden. Genome Med. 9, 34 (2017).

5. Wong, M. et al. Whole genome, transcriptome and methylome profiling enhances actionable target discovery in high-risk pediatric cancer. Nat. Med. 26, 1742–1753 (2020).

6. Pleasance, E. et al. Whole-genome and transcriptome analysis enhances precision cancer treatment options. Ann. Oncol. Off. J. Eur. Soc. Med. Oncol. 33, 939–949 (2022).

7. van der Velden, D. L. et al. The Drug Rediscovery protocol facilitates the expanded use of existing anticancer drugs. Nature 574, 127–131 (2019).

8. Shukla, N. et al. Feasibility of whole genome and transcriptome profiling in pediatric and young adult cancers. Nat. Commun. 13, 2485 (2022).

9. Cuppen, E. et al. Implementation of Whole-Genome and Transcriptome Sequencing Into Clinical Cancer Care. *JCO Precis*. Oncol. 6, e2200245 (2022).

10. Logsdon, G. A., Vollger, M. R. & Eichler, E. E. Long-read human genome sequencing and its applications. Nat. Rev. Genet. 21, 597–614 (2020).

11. Ebert, P. et al. Haplotype-resolved diverse human genomes and integrated analysis of structural variation. Science 372, eabf7117 (2021).

12. Yuen, Z. W.-S. et al. Systematic benchmarking of tools for CpG methylation detection from nanopore sequencing. Nat. Commun. 12, 3438 (2021).

13. Simpson, J. T. et al. Detecting DNA cytosine methylation using nanopore sequencing. Nat. Methods 14, 407–410 (2017).

14. Berdasco, M. & Esteller, M. Clinical epigenetics: seizing opportunities for translation. Nat. Rev. Genet. 20, 109–127 (2019).

15. Jones, P. A., Issa, J.-P. J. & Baylin, S. Targeting the cancer epigenome for therapy. Nat. Rev. Genet. 17, 630–641 (2016).

16. Rausch, T. et al. Long-read sequencing of diagnosis and post-therapy medulloblastoma reveals complex rearrangement patterns and epigenetic signatures. Cell Genomics 3, 100281 (2023).

17. Fujimoto, A. et al. Whole-genome sequencing with long reads reveals complex structure and origin of structural variation in human genetic variations and somatic mutations in cancer. Genome Med. 13, 65 (2021).

18. Aganezov, S. et al. Comprehensive analysis of structural variants in breast cancer genomes using single-molecule sequencing. Genome Res. 30, 1258–1273 (2020).

19. Wang, Y., Zhao, Y., Bollas, A., Wang, Y. & Au, K. F. Nanopore sequencing technology, bioinformatics and applications. Nat. Biotechnol. 39, 1348–1365 (2021).

20. Shiraishi, Y. et al. Precise characterization of somatic complex structural variations from tumor/control paired long-read sequencing data with nanomonsv. Nucleic Acids Res. 51, e74 (2023).

21. Chu, J. et al. BioBloom tools: fast, accurate and memory-efficient host species sequence screening using bloom filters. Bioinforma. Oxf. Engl. 30, 3402–3404 (2014).

22. Tsang, E. S. et al. Uncovering Clinically Relevant Gene Fusions with Integrated Genomic and Transcriptomic Profiling of Metastatic Cancers. Clin. Cancer Res. Off. J. Am. Assoc. Cancer Res. 27, 522–531 (2021).

23. Xu, L. et al. Long-read sequencing identifies novel structural variations in colorectal cancer. PLOS Genet. 19, e1010514 (2023).

24. Hadi, K. et al. Distinct Classes of Complex Structural Variation Uncovered across Thousands of Cancer Genome Graphs. Cell 183, 197–210.e32 (2020).

25. Cleary, S. & Seoighe, C. Perspectives on Allele-Specific Expression. Annu. Rev. Biomed. Data Sci. 4, 101–122 (2021).

26. Robles-Espinoza, C. D., Mohammadi, P., Bonilla, X. & Gutierrez-Arcelus, M. Allele-specific expression: applications in cancer and technical considerations. Curr. Opin. Genet. Dev. 66, 10–19 (2021).

27. Deonovic, B., Wang, Y., Weirather, J., Wang, X.-J. & Au, K. F. IDP-ASE: haplotyping and quantifying allele-specific expression at the gene and gene isoform level by hybrid sequencing. Nucleic Acids Res. 45, e32 (2017).

28. Sen, A., Huo, Y., Elster, J., Zage, P. E. & McVicker, G. Allele-specific expression reveals genes with recurrent cis-regulatory alterations in high-risk neuroblastoma. Genome Biol. 23, 71 (2022).

29. Clayton, E. A. et al. Tumor suppressor genes and allele-specific expression: mechanisms and significance. Oncotarget 11, 462–479 (2020).

30. Paydas, S. Prognostic Importance of DUSP22 (Dual Specificity Phosphatase 22) Gene Expression in Low-Grade Lymphomas. *Eurasian J*. Med. Oncol. (2022) doi:10.14744/ejmo.2021.59080.

31. Pedersen, M. B. et al. DUSP22 and TP63 rearrangements predict outcome of ALK-negative anaplastic large cell lymphoma: a Danish cohort study. Blood 130, 554–557 (2017).

32. Lin, C.-Y. et al. Allele-specific expression in a family quartet with autism reveals mono-to-biallelic switch and novel transcriptional processes of autism susceptibility genes. Sci. Rep. 8, 4277 (2018).

33. Stelzer, Y. et al. Differentiation of human parthenogenetic pluripotent stem cells reveals multiple tissue- and isoform-specific imprinted transcripts. Cell Rep. 11, 308–320 (2015).

34. Castel, S. E. et al. A vast resource of allelic expression data spanning human tissues. Genome Biol. 21, 234 (2020).

35. Amarasinghe, S. L. et al. Opportunities and challenges in long-read sequencing data analysis. Genome Biol. 21, 30 (2020).

36. Dawson, M. A. & Kouzarides, T. Cancer Epigenetics: From Mechanism to Therapy. Cell 150, 12–27 (2012).

37. Rasmussen, K. D. & Helin, K. Role of TET enzymes in DNA methylation, development, and cancer. Genes Dev. 30, 733–750 (2016).

38. Wu, H. et al. Dual functions of Tet1 in transcriptional regulation in mouse embryonic stem cells. Nature 473, 389–393 (2011).

39. Ko, M. et al. Modulation of TET2 expression and 5-methylcytosine oxidation by the CXXC domain protein IDAX. Nature 497, 122–126 (2013).

40. Wild, L. & Flanagan, J. M. Genome-wide hypomethylation in cancer may be a passive consequence of transformation. Biochim. Biophys. Acta 1806, 50–57 (2010).

41. Zhang, W. et al. Global DNA Hypomethylation in Epithelial Ovarian Cancer: Passive Demethylation and Association with Genomic Instability. Cancers 12, 764 (2020).

42. Zhou, W. et al. DNA methylation loss in late-replicating domains is linked to mitotic cell division. Nat. Genet. 50, 591–602 (2018).

43. Pratt, D., Penas-Prado, M. & Gilbert, M. R. Clinical impact of molecular profiling in rare brain tumors. Curr. Opin. Neurol. 36, 579–586 (2023).

44. Yang, Y. et al. Hierarchical classification-based pan-cancer methylation analysis to classify primary cancer. BMC Bioinformatics 24, 465 (2023).

45. Tessier-Cloutier, B. et al. The impact of whole genome and transcriptome analysis (WGTA) on predictive biomarker discovery and diagnostic accuracy of advanced malignancies. J. Pathol. Clin. Res. 8, 395–407 (2022).

46. Patterson, M. et al. WhatsHap: Weighted Haplotype Assembly for Future-Generation Sequencing Reads. J. Comput. Biol. J. Comput. Mol. Cell Biol. 22, 498–509 (2015).

47. Ibáñez, C. F. Structure and physiology of the RET receptor tyrosine kinase. Cold Spring Harb. Perspect. Biol. 5, a009134 (2013).

48. Chen, Z. et al. Comprehensive Analysis Revealed that CDKN2A is a Biomarker for Immune Infiltrates in Multiple Cancers. Front. Cell Dev. Biol. 9, 808208 (2021).

49. Creeden, J. F. et al. Homologous recombination proficiency in ovarian and breast cancer patients. BMC Cancer 21, 1154 (2021).

50. Hansmann, T. et al. Constitutive promoter methylation of BRCA1 and RAD51C in patients with familial ovarian cancer and early-onset sporadic breast cancer. Hum. Mol. Genet. 21, 4669–4679 (2012).

51. Esteller, M. et al. Promoter hypermethylation and BRCA1 inactivation in sporadic breast and ovarian tumors. J. Natl. Cancer Inst. 92, 564–569 (2000).

52. Suter, C. M., Martin, D. I. K. & Ward, R. L. Germline epimutation of MLH1 in individuals with multiple cancers. Nat. Genet. 36, 497–501 (2004).

53. Deshpande, V. et al. Exploring the landscape of focal amplifications in cancer using AmpliconArchitect. Nat. Commun. 10, 392 (2019).

54. Chua, Y. L. et al. The NRG1 gene is frequently silenced by methylation in breast cancers and is a strong candidate for the 8p tumour suppressor gene. Oncogene 28, 4041–4052 (2009).

55. Huang, H.-E. et al. A Recurrent Chromosome Breakpoint in Breast Cancer at the *NRG1* / *Neuregulin 1* / *Heregulin* Gene. Cancer Res. 64, 6840–6844 (2004).

56. Lee, C. et al. Epigenetic regulation of Neuregulin 1 promotes breast cancer progression associated to hyperglycemia. Nat. Commun. 14, 439 (2023).

57. Yang, L. et al. NRG1-dependent activation of HER3 induces primary resistance to trastuzumab in HER2-overexpressing breast cancer cells. Int. J. Oncol. 51, 1553–1562 (2017).

58. Akagi, K. et al. Intratumoral Heterogeneity and Clonal Evolution Induced by HPV Integration. Cancer Discov. 13, 910–927 (2023).

59. Gagliardi, A. et al. Analysis of Ugandan cervical carcinomas identifies human papillomavirus clade-specific epigenome and transcriptome landscapes. Nat. Genet. 52, 800–810 (2020).

60. Gordeeva, O. Cancer-testis antigens: Unique cancer stem cell biomarkers and targets for cancer therapy. Semin. Cancer Biol. 53, 75–89 (2018).

61. Bae, J. M. et al. Identification of tissue of origin in cancer of unknown primary using a targeted bisulfite sequencing panel. Epigenomics 14, 615–628 (2022).

62. Nagle, C. M. et al. Endometrial cancer risk and survival by tumor MMR status. J. Gynecol. Oncol. 29, e39 (2018).

63. Wang, Q. et al. Gene body methylation in cancer: molecular mechanisms and clinical applications. Clin. Epigenetics 14, 154 (2022).

64. Jeziorska, D. M. et al. DNA methylation of intragenic CpG islands depends on their transcriptional activity during differentiation and disease. Proc. Natl. Acad. Sci. U. S. A. 114, E7526–E7535 (2017).

65. Amante, S. M. et al. Transcription of intragenic CpG islands influences spatiotemporal host gene pre-mRNA processing. Nucleic Acids Res. 48, 8349–8359 (2020).

66. Wang, Y.-W. et al. ITPKA Gene Body Methylation Regulates Gene Expression and Serves as an Early Diagnostic Marker in Lung and Other Cancers. J. Thorac. Oncol. Off. Publ. Int. Assoc. Study Lung Cancer 11, 1469–1481 (2016).

67. Wong, C. H. et al. The establishment of CDK9/RNA PolII/H3K4me3/DNA methylation feedback promotes HOTAIR expression by RNA elongation enhancement in cancer. Mol. Ther. J. Am. Soc. Gene Ther. 30, 1597–1609 (2022).

68. Nguyen, L., W M Martens, J., Van Hoeck, A. & Cuppen, E. Pan-cancer landscape of homologous recombination deficiency. Nat. Commun. 11, 5584 (2020).

69. McGrail, D. J. et al. Widespread BRCA1/2-independent homologous recombination defects are caused by alterations in RNA-binding proteins. Cell Rep. Med. 4, 101255 (2023).

70. Zhao, E. Y. et al. Homologous Recombination Deficiency and Platinum-Based Therapy Outcomes in Advanced Breast Cancer. Clin. Cancer Res. Off. J. Am. Assoc. Cancer Res. 23, 7521–7530 (2017).

71. O’Malley, D. M., Krivak, T. C., Kabil, N., Munley, J. & Moore, K. N. PARP Inhibitors in Ovarian Cancer: A Review. Target. Oncol. 18, 471–503 (2023).

72. Laskin, J. et al. NRG1 fusion-driven tumors: biology, detection, and the therapeutic role of afatinib and other ErbB-targeting agents. Ann. Oncol. Off. J. Eur. Soc. Med. Oncol. 31, 1693–1703 (2020).

73. Hung, K. L. et al. Targeted profiling of human extrachromosomal DNA by CRISPR-CATCH. Nat. Genet. 54, 1746–1754 (2022).

74. Gorzynski, J. E. et al. Ultrarapid Nanopore Genome Sequencing in a Critical Care Setting. N. Engl. J. Med. 386, 700–702 (2022).

75. Zheng, Z. et al. Symphonizing pileup and full-alignment for deep learning-based long-read variant calling. Nat. Comput. Sci. 2, 797–803 (2022).

76. Sedlazeck, F. J. et al. Accurate detection of complex structural variations using single-molecule sequencing. Nat. Methods 15, 461–468 (2018).

77. Smolka, M. et al. Detection of mosaic and population-level structural variants with Sniffles2. Nat. Biotechnol. (2024) doi:10.1038/s41587-023-02024-y.

78. Jiang, T. et al. Long-read-based human genomic structural variation detection with cuteSV. Genome Biol. 21, 189 (2020).

79. Akbari, V. et al. Megabase-scale methylation phasing using nanopore long reads and NanoMethPhase. Genome Biol. 22, 68 (2021).

80. Li, H. Minimap2: pairwise alignment for nucleotide sequences. Bioinforma. Oxf. Engl. 34, 3094–3100 (2018).

81. Kim, S. et al. Strelka2: fast and accurate calling of germline and somatic variants. Nat. Methods 15, 591–594 (2018).

82. Simpson, J. T. et al. ABySS: a parallel assembler for short read sequence data. Genome Res. 19, 1117–1123 (2009).

83. Birol, I. et al. De novo transcriptome assembly with ABySS. Bioinforma. Oxf. Engl. 25, 2872–2877 (2009).

84. Chen, X., et al. Manta: rapid detection of structural variants and indels for germline and cancer sequencing applications. Bioinforma. Oxf. Engl. 32, 1220–1222 (2016).

85. Rausch, T., et al. DELLY: structural variant discovery by integrated paired-end and split-read analysis. Bioinforma. Oxf. Engl. 28, i333–i339 (2012).

86. Reisle, C. et al. MAVIS: merging, annotation, validation, and illustration of structural variants. Bioinformatics 35, 515–517 (2019).

87. Cingolani, P. et al. A program for annotating and predicting the effects of single nucleotide polymorphisms, SnpEff: SNPs in the genome of Drosophila melanogaster strain w1118; iso-2; iso-3. Fly (Austin) 6, 80–92 (2012).

88. Cunningham, F. et al. Ensembl 2022. Nucleic Acids Res. 50, D988–D995 (2022).

89. Niu, B. et al. MSIsensor: microsatellite instability detection using paired tumor-normal sequence data. Bioinforma. Oxf. Engl. 30, 1015–1016 (2014).

90. Dobin, A. et al. STAR: ultrafast universal RNA-seq aligner. Bioinforma. Oxf. Engl. 29, 15–21 (2013).

91. Li, B. & Dewey, C. N. RSEM: accurate transcript quantification from RNA-Seq data with or without a reference genome. BMC Bioinformatics 12, 323 (2011).

92. MacDonald, J. R., Ziman, R., Yuen, R. K. C., Feuk, L. & Scherer, S. W. The Database of Genomic Variants: a curated collection of structural variation in the human genome. Nucleic Acids Res. 42, D986–992 (2014).

93. Smit, A., Hubley, R. & Green, P. RepeatMasker Open. (2013).

94. Chakravarty, D. et al. OncoKB: A Precision Oncology Knowledge Base. JCO Precis. Oncol. 2017, PO.17.00011 (2017).

95. Cortés-Ciriano, I. et al. Comprehensive analysis of chromothripsis in 2,658 human cancers using whole-genome sequencing. Nat. Genet. 52, 331–341 (2020).

96. Porter, V. L., et al. Genomic Structures and Regulation Patterns at HPV Integration Sites in Cervical Cancer. http://biorxiv.org/lookup/doi/10.1101/2023.11.04.564800<x> (2023) doi:10.1101/2023.11.04.564800.

97. Mayba, O. et al. MBASED: allele-specific expression detection in cancer tissues and cell lines. Genome Biol. 15, 405 (2014).

98. Grant, C. E., Bailey, T. L. & Noble, W. S. FIMO: scanning for occurrences of a given motif. Bioinforma. Oxf. Engl. 27, 1017–1018 (2011).

99. Bernstein, B. E. et al. The NIH Roadmap Epigenomics Mapping Consortium. Nat. Biotechnol. 28, 1045–1048 (2010).

100. ENCODE Project Consortium. An integrated encyclopedia of DNA elements in the human genome. Nature 489, 57–74 (2012).

101. Loyfer, N. et al. A DNA methylation atlas of normal human cell types. Nature 613, 355–364 (2023).

102. Karolchik, D. et al. The UCSC Table Browser data retrieval tool. Nucleic Acids Res. 32, D493–496 (2004).

103. Wang, R., Nambiar, R., Zheng, D. & Tian, B. PolyA_DB 3 catalogs cleavage and polyadenylation sites identified by deep sequencing in multiple genomes. Nucleic Acids Res. 46, D315–D319 (2018).

104. Fishilevich, S. et al. GeneHancer: genome-wide integration of enhancers and target genes in GeneCards. Database J. Biol. Databases Curation 2017, bax028 (2017).

105. Park, Y. & Wu, H. Differential methylation analysis for BS-seq data under general experimental design. Bioinforma. Oxf. Engl. 32, 1446–1453 (2016).

106. Kuleshov, M. V. et al. Enrichr: a comprehensive gene set enrichment analysis web server 2016 update. Nucleic Acids Res. 44, W90–97 (2016).

107. Davies, H. et al. HRDetect is a predictor of BRCA1 and BRCA2 deficiency based on mutational signatures. Nat. Med. 23, 517–525 (2017).

108. Nik-Zainal, S. et al. Landscape of somatic mutations in 560 breast cancer whole-genome sequences. Nature 534, 47–54 (2016).

109. Talevich, E., Shain, A. H., Botton, T. & Bastian, B. C. CNVkit: Genome-Wide Copy Number Detection and Visualization from Targeted DNA Sequencing. PLOS Comput. Biol. 12, e1004873 (2016).

110. Krzywinski, M. et al. Circos: An information aesthetic for comparative genomics. Genome Res. 19, 1639–1645 (2009).

111. Therneau, T. M. & Lumley, T. Package “survival.” R Top Doc. 128, 28–33 (2015).

112. Sjoberg, D., Baillie, M., Haesendonckx, S. & Treis, T. ggsurvfit: Flexible Time-to-Event Figures. (2023).

113. Grolemund, G. & Wickham, H. Dates and Times Made Easy with lubridate. J. Stat. Softw. 40, (2011).

114. Wickham, H. et al. Welcome to the Tidyverse. J. Open Source Softw. 4, 1686 (2019).

115. Robinson, J. T. Integrative genomics viewer. Nat. Biotechnol. 29, (2011).

116. Cavalcante, R. G. & Sartor, M. A. annotatr: genomic regions in context. Bioinformatics 33, 2381–2383 (2017).

117. Pedersen, T. patchwork: The Composer of Plots. (2023).

118. Yin, T., Cook, D. & Lawrence, M. ggbio: an R package for extending the grammar of graphics for genomic data. Genome Biol. 13, R77 (2012).

119. Hothorn, T., Hornik, K., van de Wiel, M. A. & Zeileis, A. Implementing a Class of Permutation Tests: The coin Package. J Stat Sofw 28, 1–23 (2008).

