## Extended Data Figures for "Long-read sequencing of an advanced cancer cohort resolves rearrangements, unravels haplotypes, and reveals methylation landscapes"

**Extended Data Tables and Figures**

**Supplementary Table 1.** Samples in Nanopore POG cohort.
**[See Supplementary_Table_1.xlsx]**

**Supplementary Table 2**. Clinically relevant SVs alongside their corresponding types, the callers support, and their corresponding coverage.
**[See Supplementary_Table_2.xlsx]**

**Supplementary Table 3.** Summary of event counts for somatic SVs. Values are presented as median (interquartile range, 25th–75th percentile) per sample. LQ = Low Quality. HQ = High Quality.
**[See Supplementary_Table_3.xlsx]**

**Supplementary Table 4.** Insertion events involving an OncoKB gene unique to Nanopore alongside their library, the genes involved, genomic coordinates, insert size, and additional comments after manual review.
**[See Supplementary_Table_4.xlsx]**

**Supplementary Table 5.** Structural variant events predicted exclusively by Illumina or Nanopore alongside their corresponding event type, name of callers which support the event, and the corresponding long and short reads sequencing coverages. (n = 55).
**[See Supplementary_Table_5.xlsx]**

**Supplementary Table 6. Summary of genes detected on ecDNAs in cohort.** Genes detected on ecDNAs, their relative frequencies, and whether they are predicted as oncogenes.
**[See Supplementary_Table_6.xlsx]**

**Supplementary Table 7. ENCODE accessions for normal WGBS samples.**
**[See Supplementary_Table_7.xlsx]**

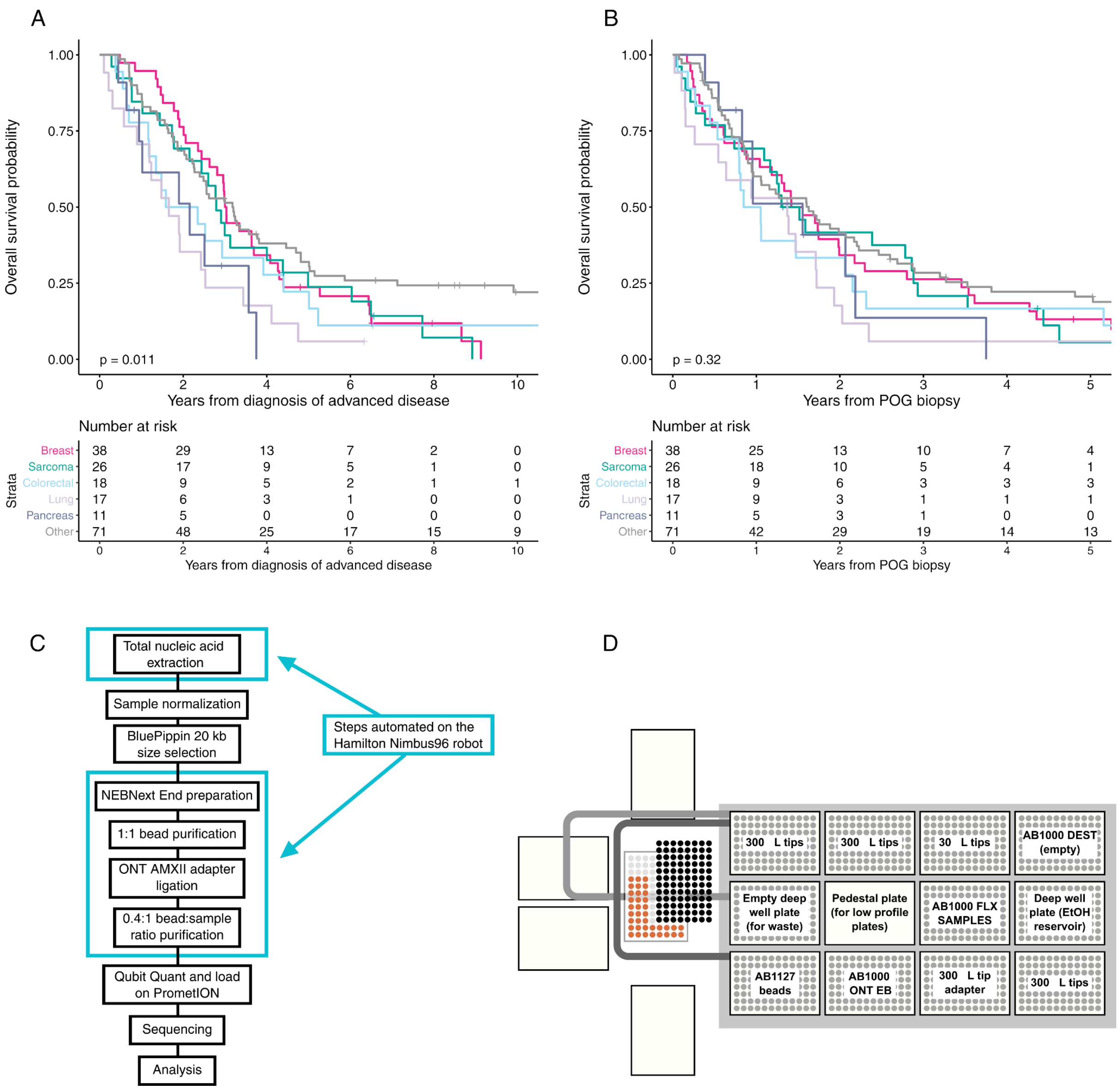


**Extended Data Figure 1. Cohort and sequencing information.** (A-B) Overall survival for Long-read POG cohort by tumour type from date of advanced disease (A) and from date of biopsy (B). (C) Workflow for Nanopore sample preparation highlighting steps automated on the Nimbus96 liquid handler. (D) Nimbus96 robot deck layout for magnetic bead purification.

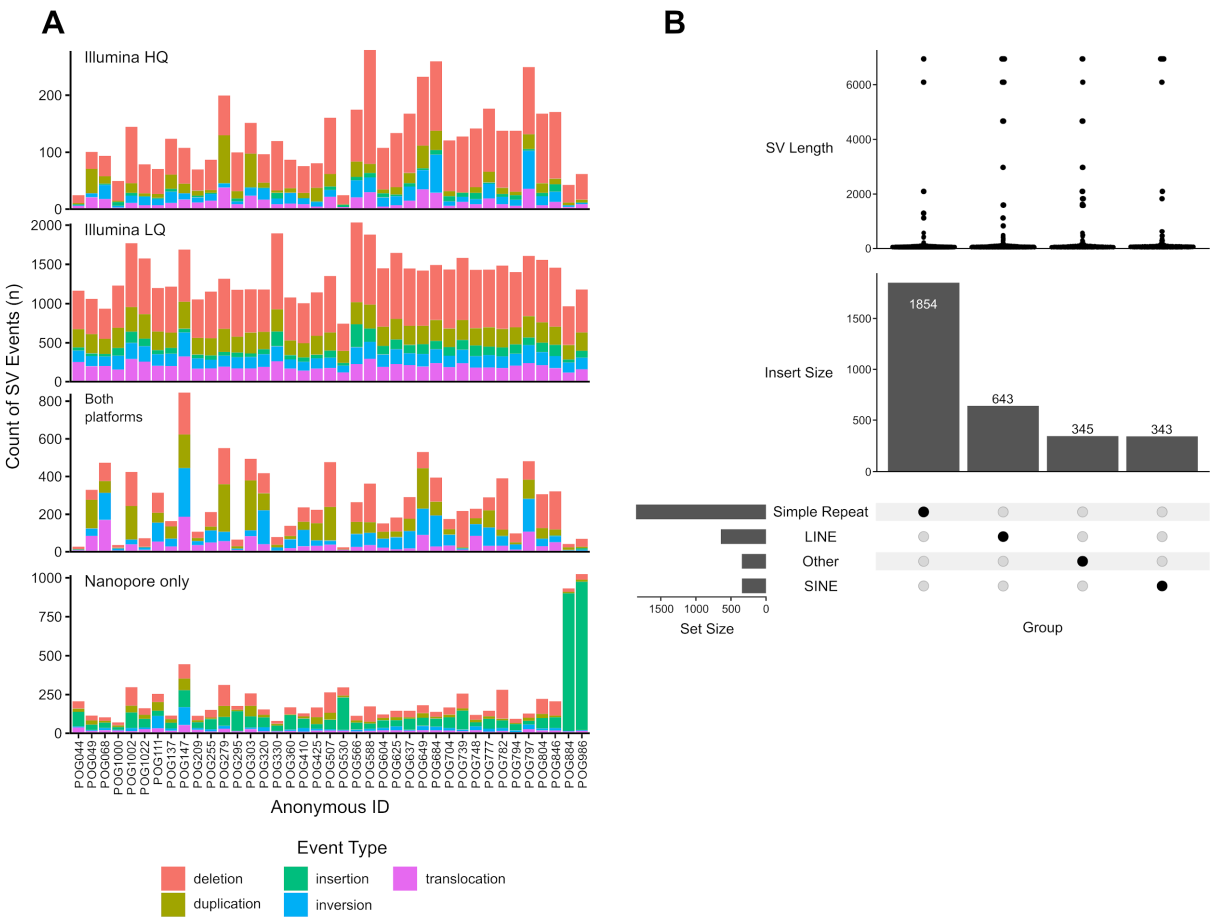


**Extended Data Figure 2. SV distributions and examples**. (A) Number of somatic SV events per sample in different call sets. POG111 and POG147 stand out as having much more inversions called in the long-read data than all other cases. POG884 and POG986 stand out as having much more insertions called. (B) Nanopore-only insertion calls in an MSI-H sample POG986, annotated with RepeatMasker. Other repetitive elements such as Alu elements within RepeatMasker are demarcated as “other”

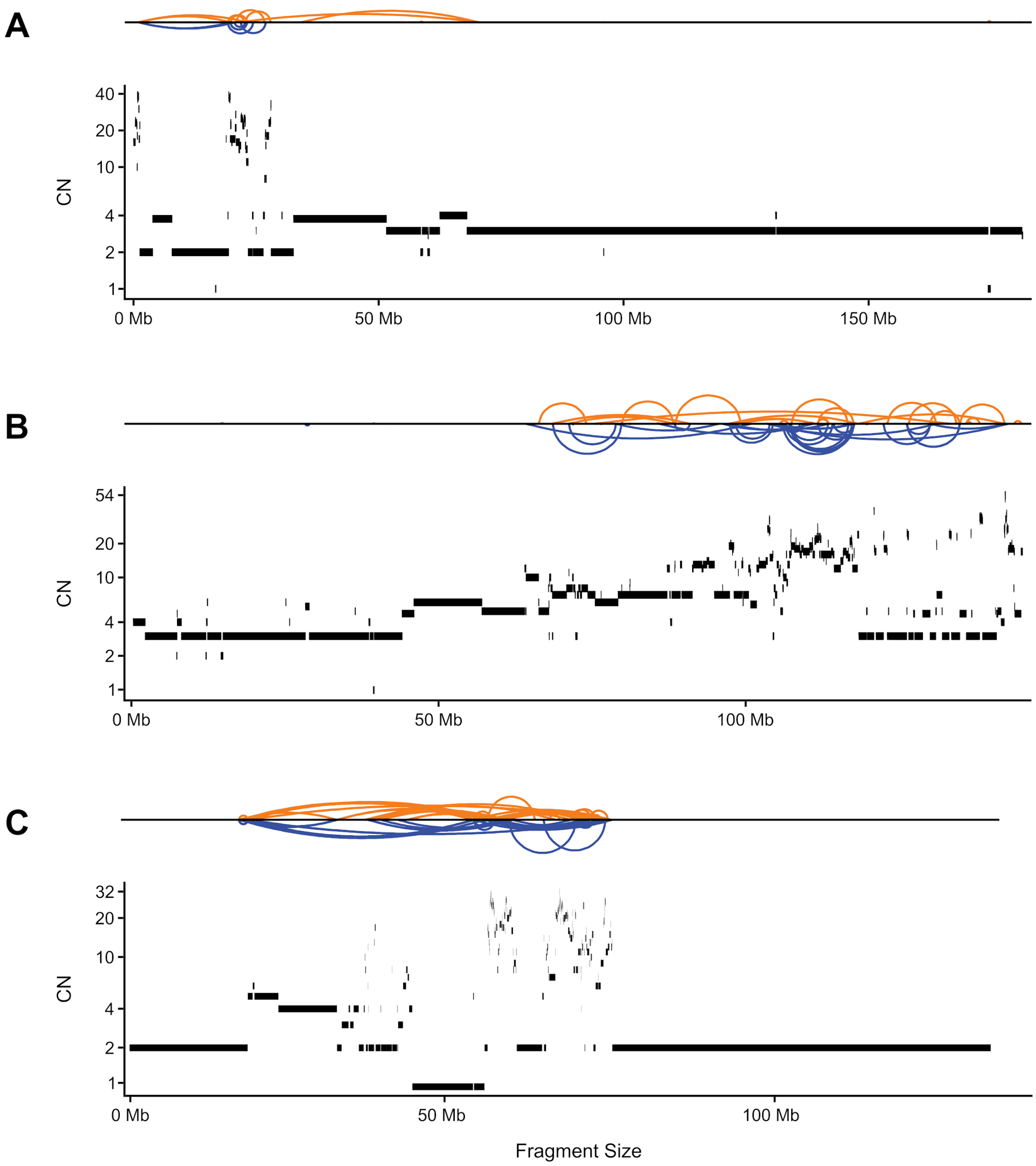


**Extended Data Figure 3. Examples of complex SVs.** (A-C) Shatterseek images of patient POG147 (A) and patient POG111 (B,C) with the 'Sniffles' structural variant profiles (top of subplots), and Ploidetect copy number calls (bottom of subplots). Duplication-like SVs are coloured blue, and deletion-like SVs are coloured orange, and highlight the tyfona-like structural variant profile of chromosome 5 (A) and chromosome 8 (B,C).

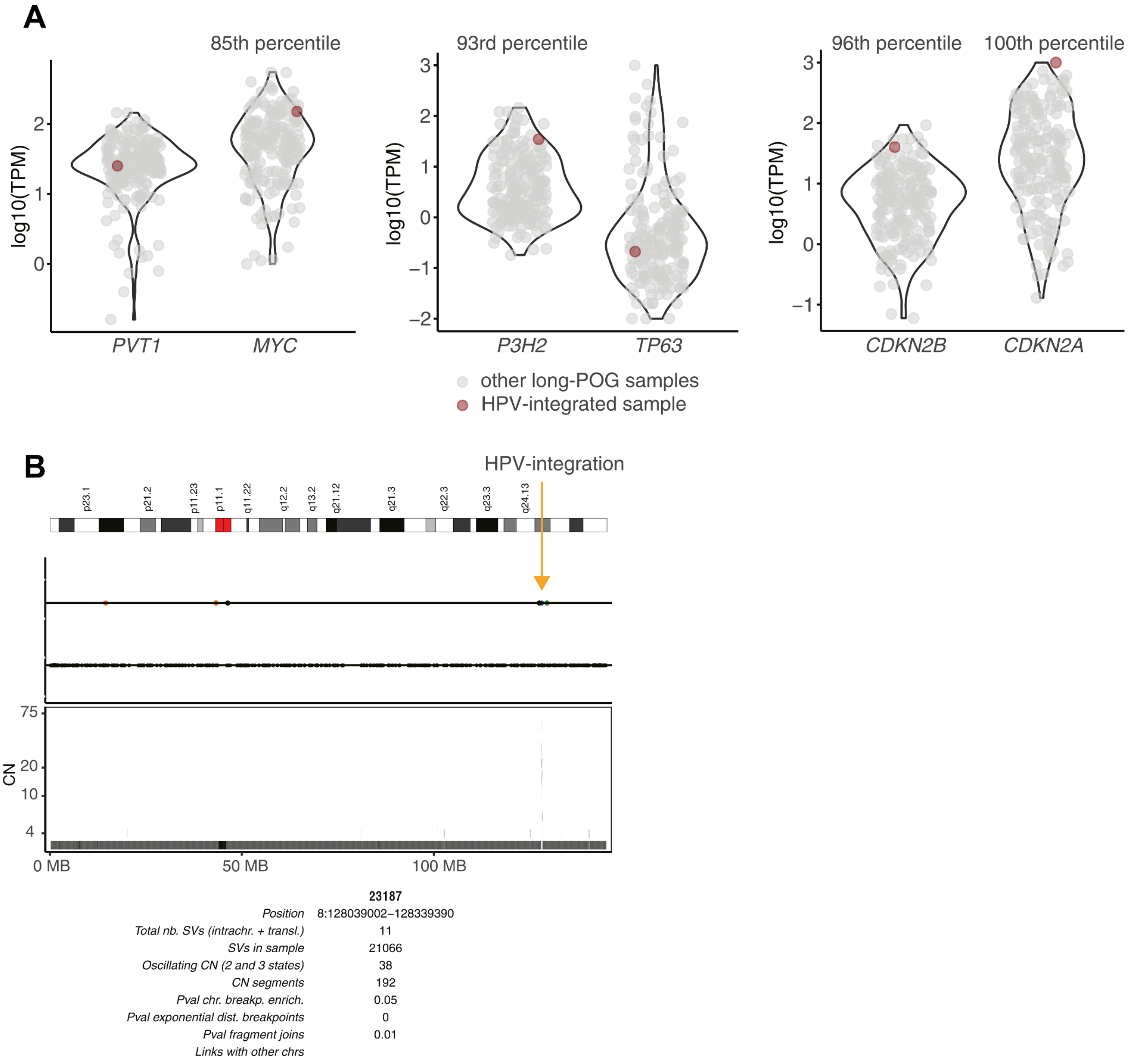


**Extended Data Figure 4. HPV integration events.** (a) Gene expression in HPV-integrated samples. (b) Focal chromothripsis-like event at HPV integration site.

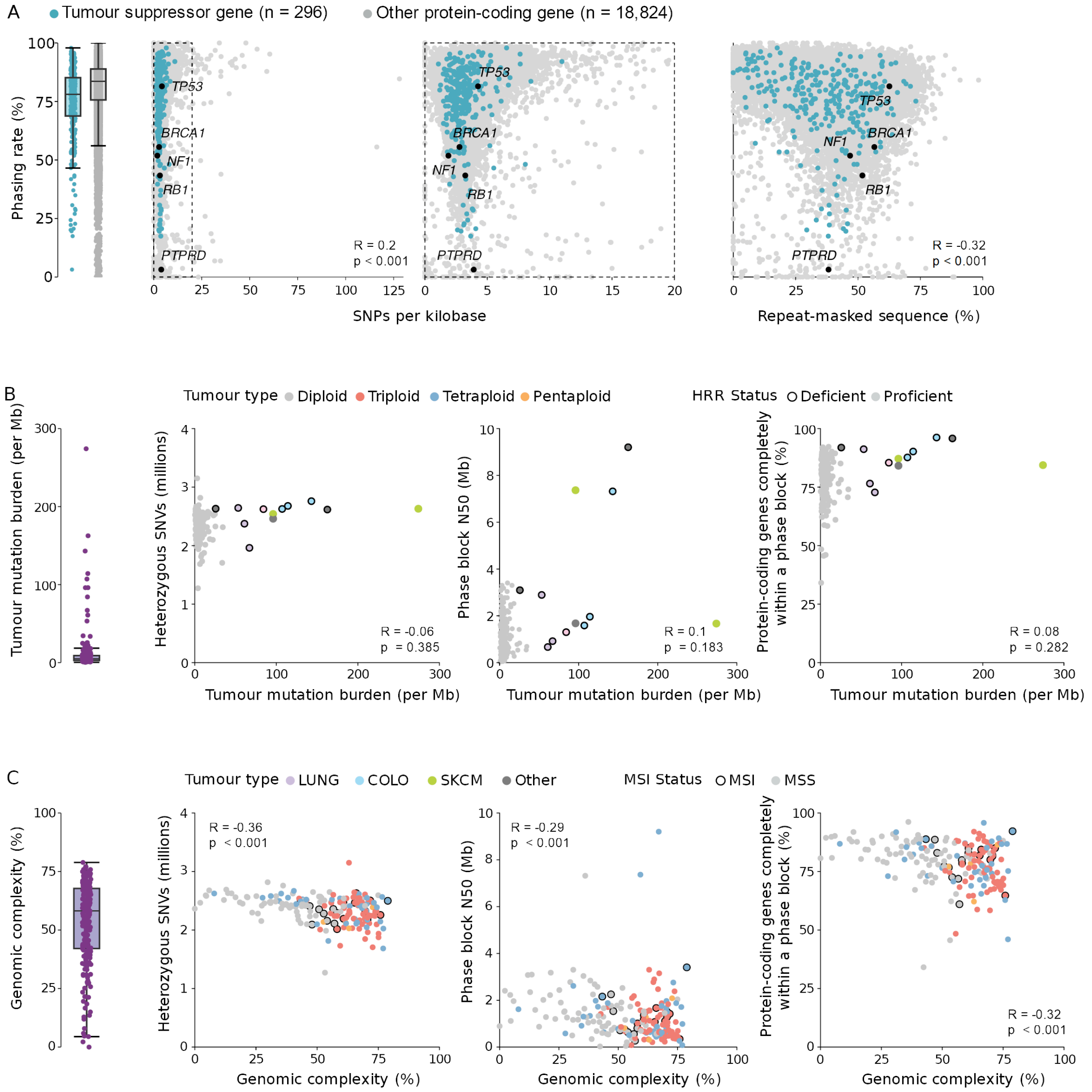


**Extended Data Figure 5. Factors affecting phasing in long-read sequenced tumours.** (A) Effect of germline factors on phasing rate per-gene. SNP density (left) and proportion of the gene body made up of repeats (right) both correlate weakly with phasing rate. (B) Effects of somatic small variants (represented by TMB) on phasing. Neither phase block N50 nor completeness of phasing were significantly correlated. (C) Effect of somatic rearrangements (measured by genomic complexity) on phasing. Both phase block N50 and completeness of phasing were correlated, slightly more so than SNP density and repeats.

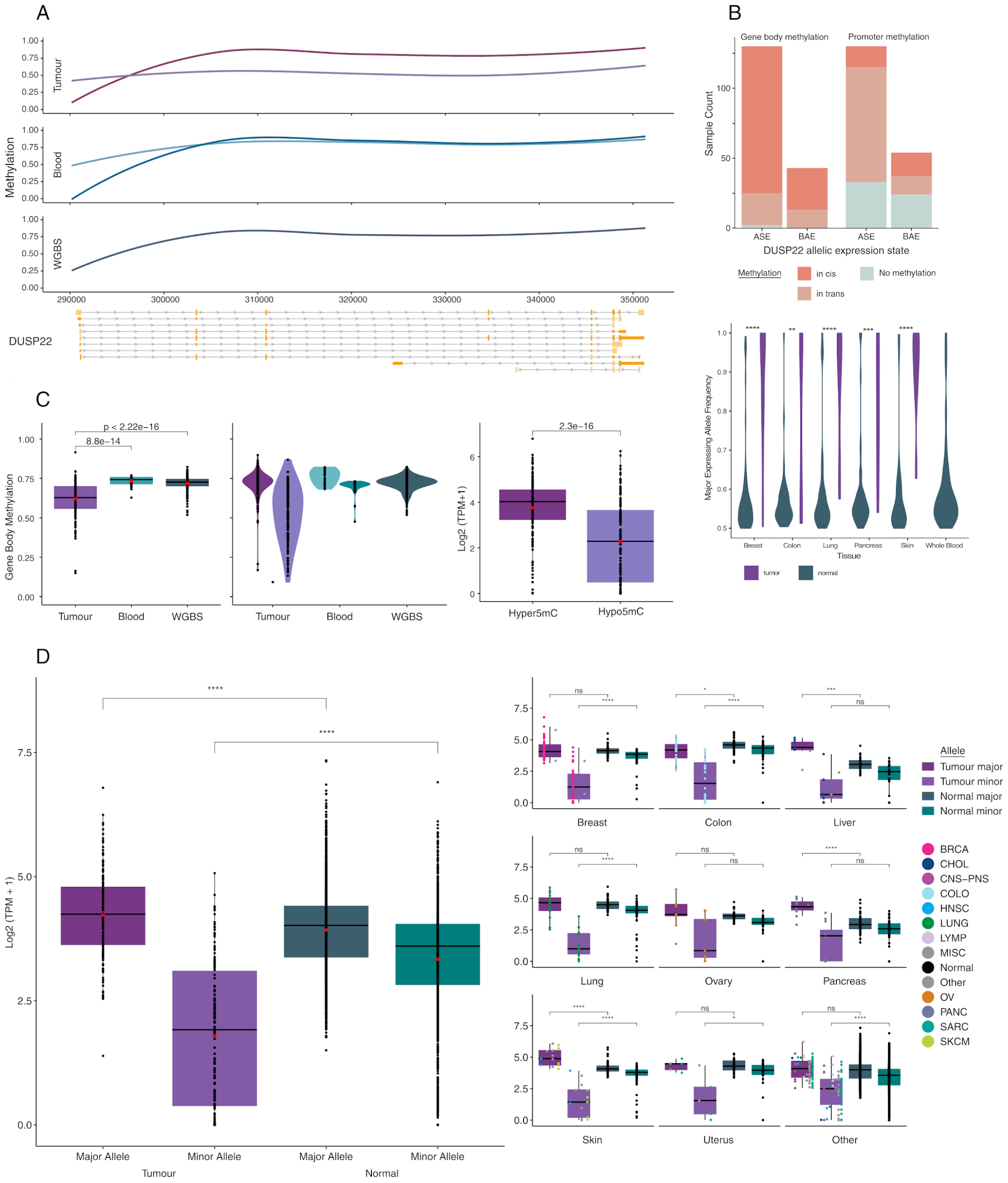


**Extended Data Figure 6. Phasing of methylation information.** (A) Allelic methylation of CpGs across *DUSP22*. For blood and tumour tracks, lighter points indicate hypomethylated alleles across the region whereas darker samples indicate hypermethylated alleles. The gene tracks for the top ten expressed transcripts as well as CpG islands are shown on the bottom. (C) *DUSP22* gene body methylation of tumour, blood and normal whole genome bisulfate sequencing samples at the sample and allelic level respectively (left two figures). Expression of alleles showing gene body hypermethylation in comparison to those showing hypomethylation (right). (D) Allelic expression of *DUSP22* for the major and minor allele of tumor samples compared to GTEx samples with more than 20 reads to determine major allele frequency. Allelic expression is shown across tissues of origin for cancers and normals and the color of each point indicates the cancer type. Wicoxon rank-sum test p-values and significance labels are shown above plots (*P*<0.0001: ****, *P*<0.001: ***, *P*>0.05: ns). All samples with loss of heterozygosity or deletions in this gene were filtered out.

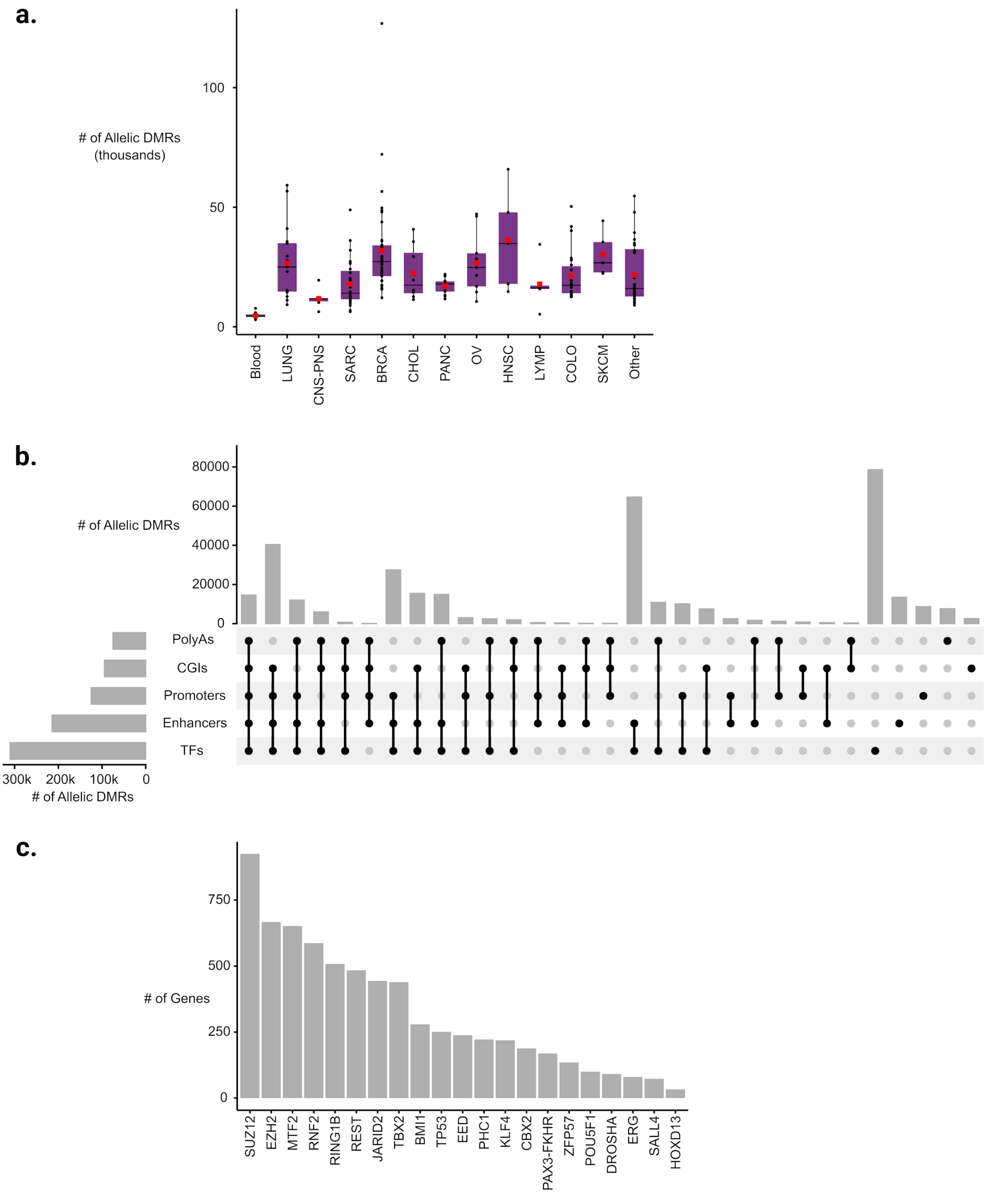


**Extended Data Figure 7. Allelic DMRs.** (a) Number of allelic DMRs in matched blood and tumour type samples. (b) Upset plot demonstrating mapping of tumour-specific allelic DMRs to different genomic regions. (c) Recurring transcription factor (TF) binding sites with tumour-specific allelic DMR. The plot represents the top 30 TFs with the highest proportion of their binding sites recurrently overlapped to tumour-specific allelic DMR.

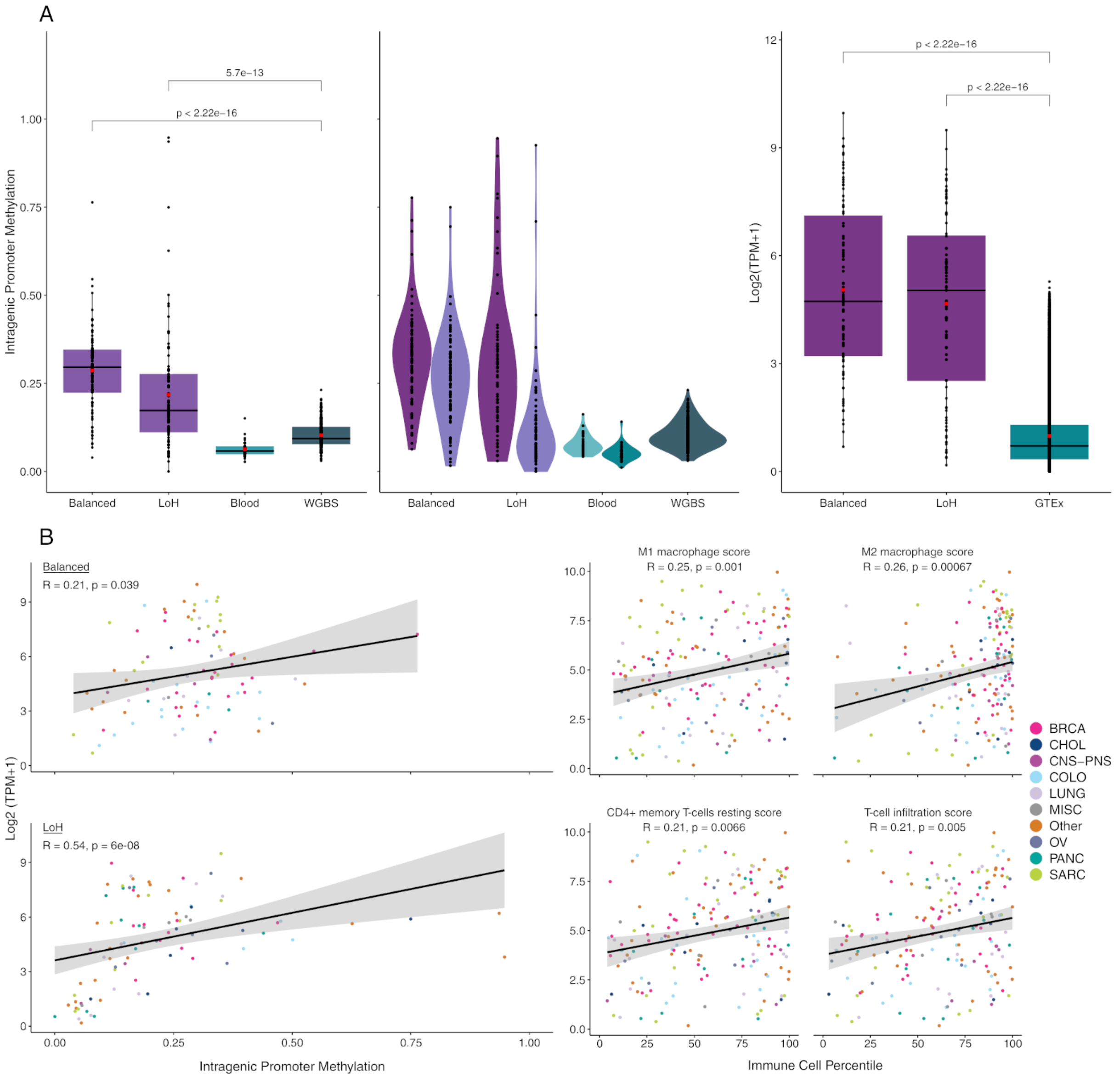


**Extended Data Figure 8. Methylation of *CDKN2A*.** (a) The left two figures show *CDKN2A* intragenic promoter methylation of balanced tumour, loss of heterozygosity tumour, blood and normal reference samples at the sample and allelic level respectively. The right figure displays the expression of balanced and loss of hetrozygosity tumor samples in comparison to Genotype-Tissue Expression (GTEx) reference expression data for all tissues. Wilcoxon rank-sum test *P*-values are shown above plots for comparisons of interest. WGBS, whole genome bisulfite sequencing. (b) The left figure shows the linear relationship between *CDKN2A* intragenic promoter methylation and expression for both balanced and loss of heterozygosity samples. The right figure shows the relationship of percentile score for various immune cell populations in sample biopsies in comparison to *CDKN2A* expression. Immune cell percentile score was inferred by CIBERSORT and the spearman R value and corresponding p-value are shown in the top left corner for each plot.

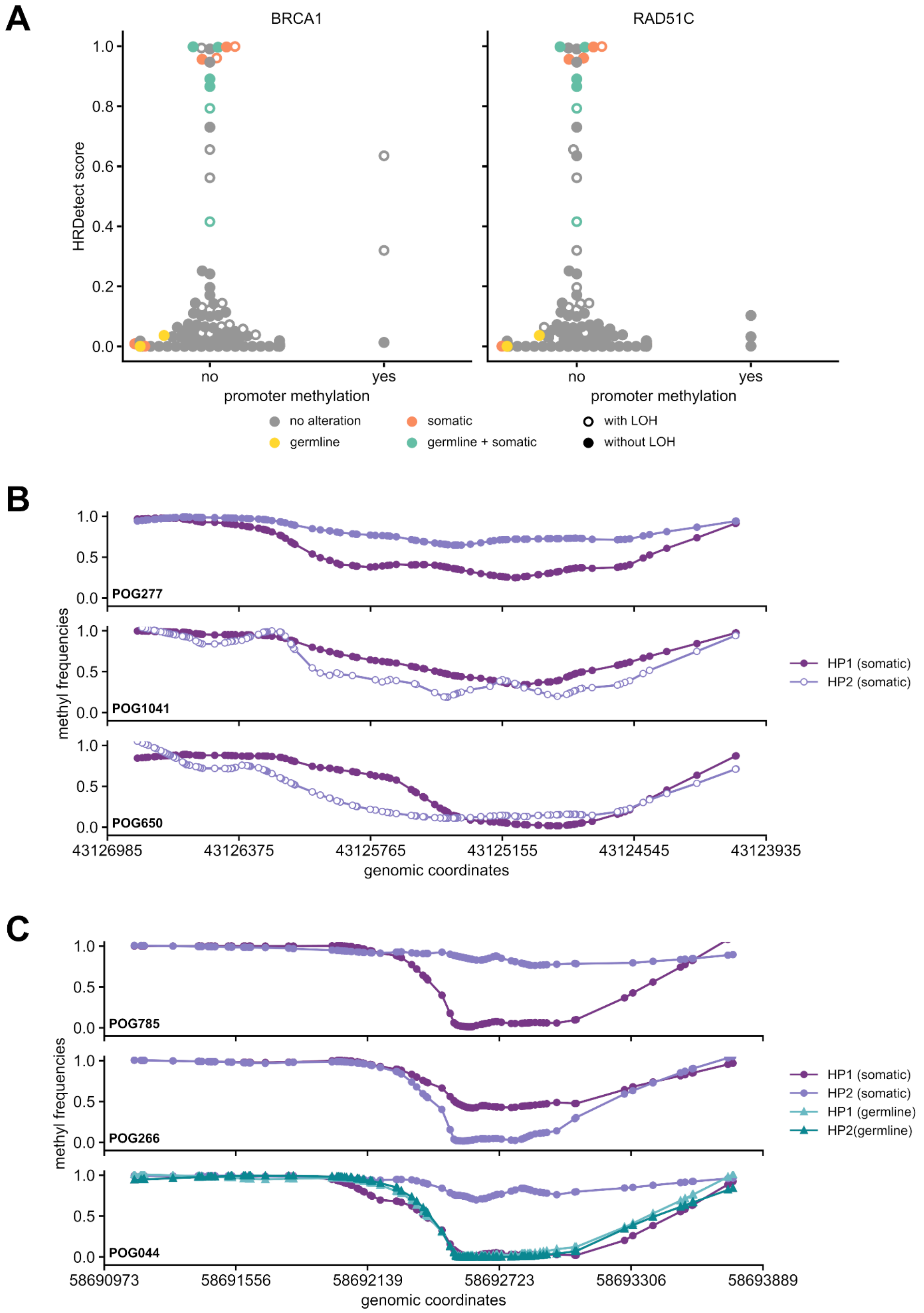


**Extended Data Figure 9. Methylation at tumour suppressor promoters.** (a) Somatic mutation status and promoter methylation in *BRCA1* and *RAD51C*. (b) Allele-specific methylation in the *BRCA1* promoter region in three cases, POG277, POG1041, POG650. (c) Allele-specific methylation in the *RAD51C* promoter region in three cases, POG785, POG266, POG044 .

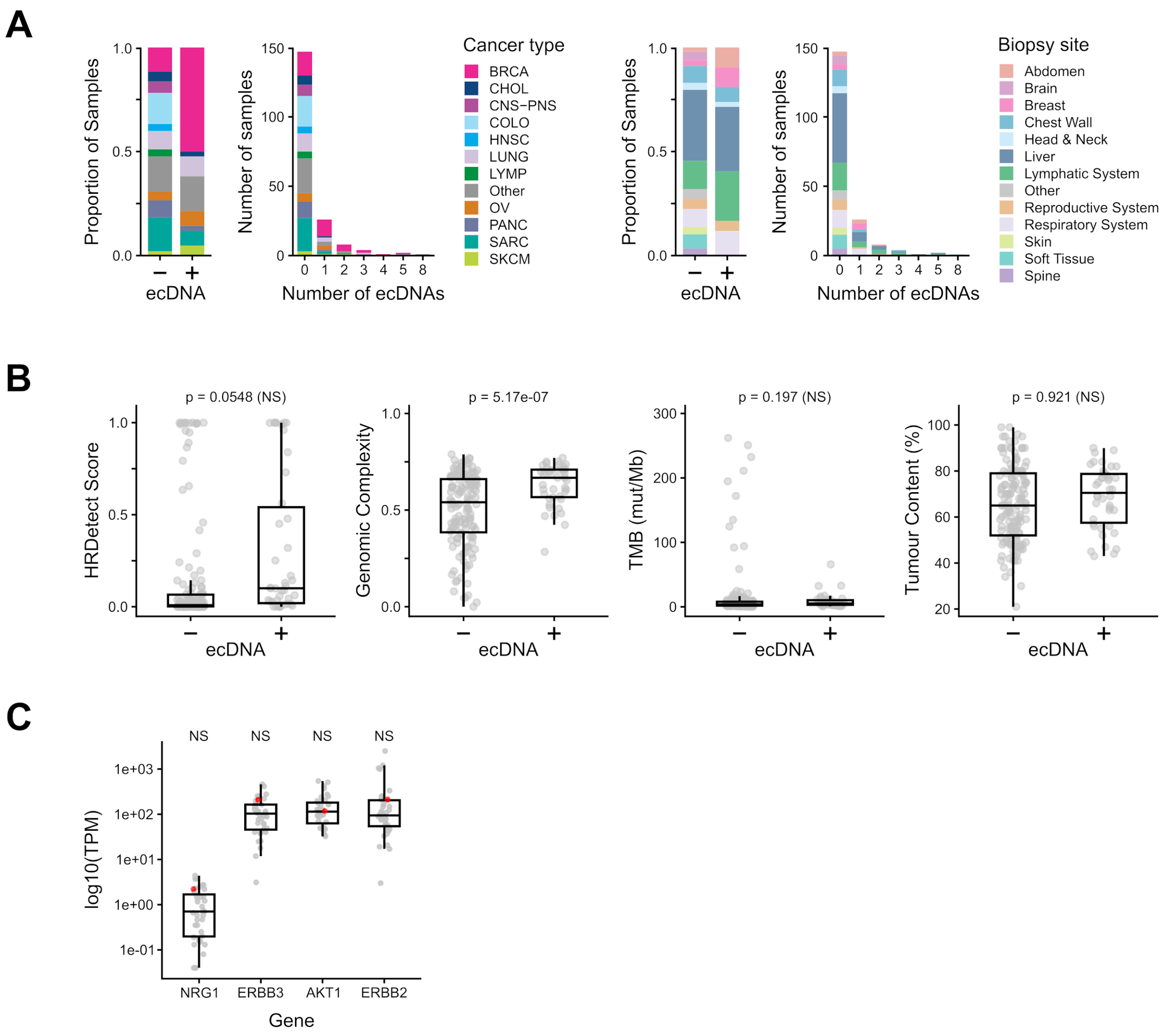


**Extended Data Figure 10. Summary of ecDNAs detected across cohort.** (A) Proportion of ecDNA+ and ecDNA- samples stratified by cancer type and biopsy site. Results obtained from running 189 samples through AmpliconArchitect, a short-read WGS ecDNA detection tool. (B) Molecular correlates of ecDNAs (n = 189). Two-sided Student's *t*-tests were used to judge significance with Bonferroni multiple testing correction. (C) Expression in transcripts per million (TPM) for *NRG1* pathway genes for ecDNA-containing breast cancer sample shown in red (*n*=1) compared to other breast cancer samples in the cohort (*n*=39). Significance assessed via one-vs-all permutation tests, with Bonferroni multiple testing correction.
