## Supplementary Figures for "Long-read sequencing of an advanced cancer cohort resolves rearrangements, unravels haplotypes, and reveals methylation landscapes"

**a**

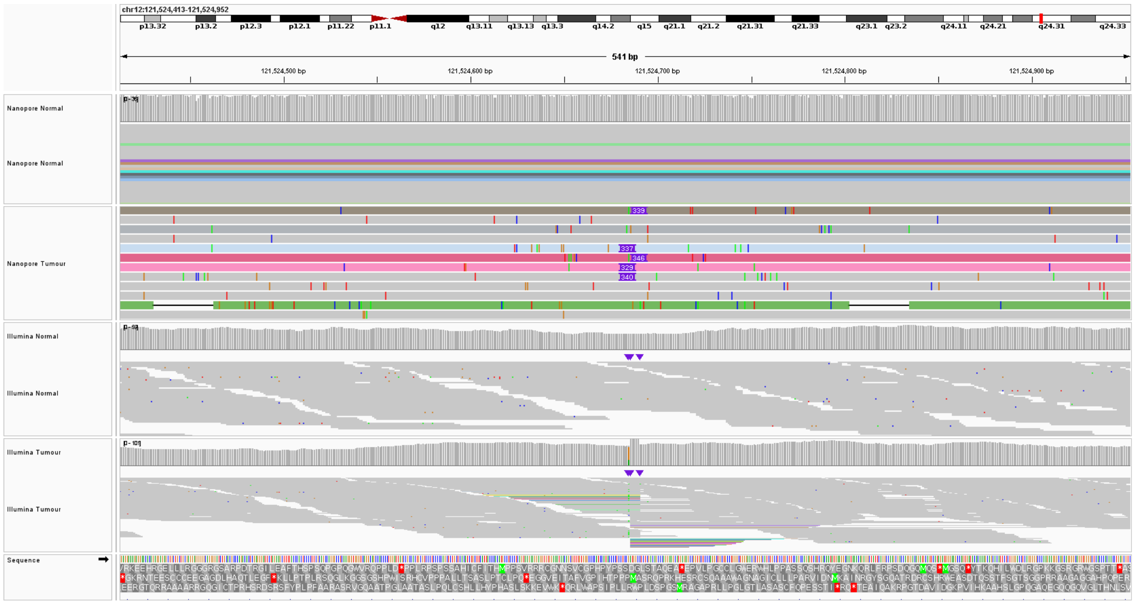


**b**

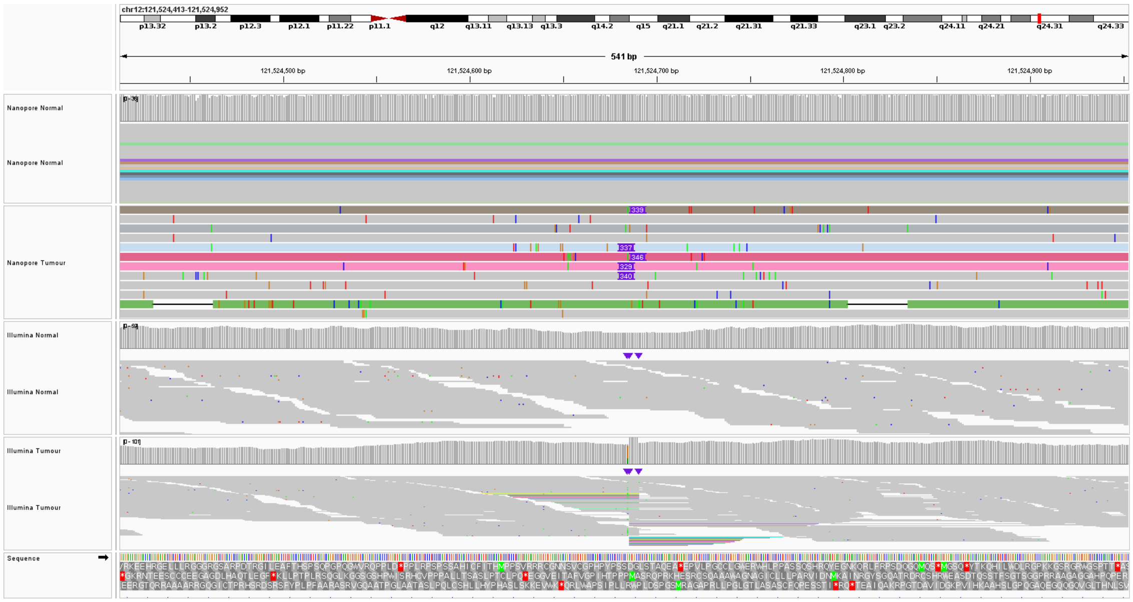


**Supplementary Figure 1. Insertion events called in long-read data but not in short-read**. (a) Intronic 90 bp insertion event on *EGFR* at chr7:55167011 by nanomonSV with no visible read based support for a 90 bp event. (b) Intronic 328 bp insertion event on *KDM2B* at chr12:121524684 called by nanomonSV miscalled by Illumina as a 111,113 bp inversion around chr12:121413571-121524684.
